# The miR-362-3p/*BCLAF1* axis regulates cisplatin sensitivity and metastatic progression in triple-negative breast cancer

**DOI:** 10.64898/2026.03.09.26347941

**Authors:** Zhaoji Liu, Catherine Wu, Madison Uyemura, Brian R Sardella, Emily K Aronson, Shizhong Ke, Erica S Massicott, Xiaohui Li, Lin Wang, Dimitra Karagkouni, Nikolas Kalavros, Ioannis S Vlachos, Felipe Batalini, Cristina S Bogsan, Jit Kong Cheong, Lihan Zhou, He Cheng, Phillip Munson, Erica L Mayer, Judy E Garber, Stuart J Schnitt, Nadine M Tung, Andrea L Kasinski, Frank J Slack, Gerburg M Wulf, Yujing J Heng

## Abstract

Platinum-based chemotherapy remains a cornerstone of treatment for triple-negative breast cancer (TNBC), yet the molecular determinants governing platinum response remain poorly defined. By leveraging the randomized Phase II INFORM trial, which compared neoadjuvant cisplatin to anthracycline-based therapy in *BRCA1/2*-mutant breast cancer—we identified miR-362-3p as a specific regulator of cisplatin sensitivity. Higher plasma miR-362-3p expression was exclusively associated with favorable clinical outcome in the cisplatin arm, with no association observed in the AC arm, decoupling platinum-specific vulnerability from general chemotherapy response. We used gain- and loss-of-function TNBC models to establish that miR-362-3p functions as a potent sensitizer to cisplatin *in vitro* and *in vivo*. Integrated TCGA analysis and experimental validation identified *BCLAF1,* a key regulator of DNA damage response, as a direct repression target of miR-362-3p. We uncovered a novel role for the miR-362-3p/*BCLAF1* axis in overcoming platinum resistance in TNBC.

## Introduction

Platinum-based chemotherapies are standard of care for various solid tumors ^1^, and remains clinically relevant for triple-negative breast cancer (TNBC) ^2–4^. The KEYNOTE-522 regimen is standard of care for stage II-III TNBC which incorporates carboplatin as a backbone of neoadjuvant treatment ^4^. Outside of the KEYNOTE-522 protocol, platinum drugs are given to breast cancer patients with advanced disease ^5^, those that have germline *BRCA1/2* mutations ^6^, or as a neoadjuvant alternative for those who cannot safely receive immunotherapy ^7^. This underscores the continued importance of understanding the biology governing platinum response in TNBC.

The INFORM trial (Translational Breast Cancer Research Consortium [TBCRC] 031) was a randomized, two-arm Phase II neoadjuvant study comparing cisplatin to an anthracycline (AC)-based regimen (doxorubicin and cyclophosphamide) in patients with germline *BRCA1/2^MUT^*, early-stage HER2-negative breast cancer (NCT01670500) ^3^. While the trial demonstrated no difference between treatment arms in rates of pathologic complete response (pCR; primary endpoint) or residual cancer burden (RCB 0/1; secondary endpoint), this well-annotated and clinically homogeneous dataset provides an unparalleled opportunity to understand the molecular determinants of general chemotherapy response from platinum sensitivity in TNBC.

MicroRNAs (miRNAs), frequently dysregulated in cancer ^8–10^, can function as dynamic regulators of oncogenic networks ^11,12^, including DNA damage response (DDR) ^13^, immune signaling ^14^, and the acquisition of therapeutic resistance ^15^. Accordingly, they have emerged as promising biomarkers of therapy response ^16^ and therapeutic targets ^17^. The identification of specific miRNA-target axes may reveal a functional layer of regulation that dictates platinum vulnerability.

By profiling pretreatment plasma samples from INFORM, we identified miR-362-3p as specifically associated with cisplatin sensitivity, but not with response to AC therapy. MiR-362-3p has been implicated as a tumor suppressor in breast, cervical, and ovarian cancers ^12,18,19^, inhibiting cellular proliferation and migration through the suppression of oncogenic targets such as *BCAR1* ^20^ and *DDX5* ^21^. Its role in the direct modulation of DDR in TNBC remains undefined.

In this study, we establish a functional role for miR-362-3p in sensitizing TNBC to platinum. Through integrated TCGA analysis and *in silico* miRNA target prediction ^22,23^, we identified and experimentally validated *BCLAF1*, a known regulator of the DDR ^24^ and apoptosis ^25^, as a direct repression target of miR-362-3p. Our findings demonstrate that the miR-362-3p/*BCLAF1* axis acts as a previously unrecognized rheostat for platinum response. This study illustrates how circulating miRNAs reflect treatment-relevant tumor stress states and provides a mechanistic blueprint for understanding the heterogeneous landscape of platinum response in TNBC.

## Results

### INFORM plasma miR-362-3p expression was exclusively associated with cisplatin response

We profiled 352 cancer-relevant miRNAs in pretreatment plasma from 100 INFORM patients. After excluding one patient who did not complete therapy and two samples that failed quality control, 97 samples were analyzed (**Fig. 1**). These 97 patients were between 24.0 to 62.8 years old (average 41.1 years old; **Table 1**). Most had a *BRCA1* mutation (71.1%), were of European ancestry (73.2%), pre-menopausal (83.5%), and had stage II disease (61.9%). About 42.3% of the patients had node involvement prior to starting treatment. Tumors were overwhelming invasive ductal carcinoma (92.8%), mostly grade 3 (78.4%) and triple negative (defined as <1% immunopositivity for estrogen receptor (ER) and progesterone receptor (PR); 66.0%). pCR and RCB 0/1 were achieved in 22.7% and 41.2% of the cases, respectively.

**Fig. 1.**
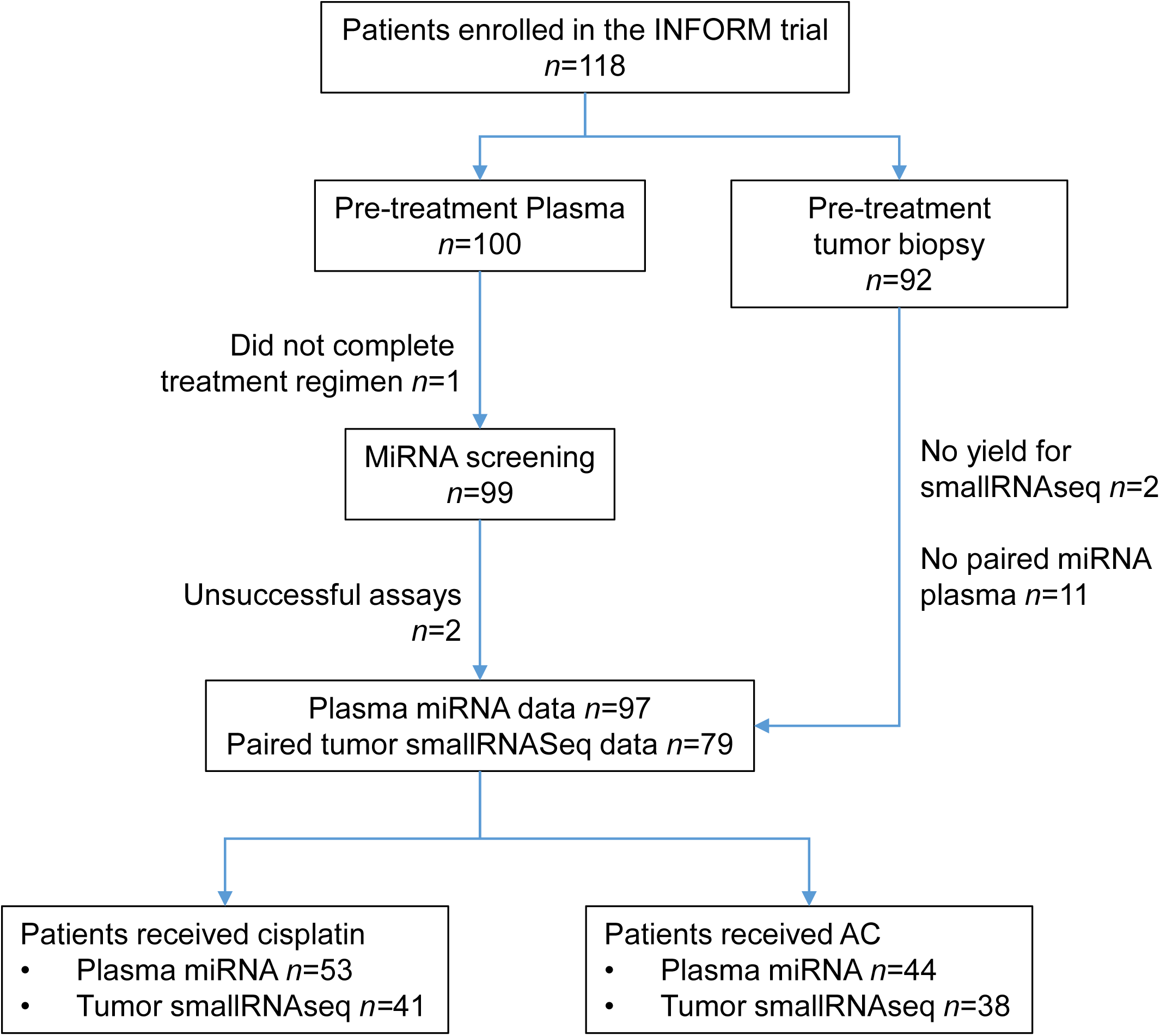
Flow diagram of INFORM patients and molecular profiling.

**Table 1.** Demographic and tumor characteristics of the INFORM participants, stratified by treatment.

|  |  | Overall | Cisplatin | AC |
| --- | --- | --- | --- | --- |
| *n* |  | 97 | 53 | 44 |
| Age, years, mean (SD) |  | 41.1 (8.9) | 39.7 (8.3) | 42.8 (9.4) |
| Age, years (%) | <40 | 45 (46.4) | 28 (52.8) | 17 (38.6) |
|  | ≥40 | 52 (53.6) | 25 (47.2) | 27 (61.4) |
| *BRCA*<sup>MUT</sup> status, *n* (%) | *BRCA1* | 69 (71.1) | 40 (75.5) | 29 (65.9) |
|  | *BRCA2* | 28 (28.9) | 13 (24.5) | 15 (34.1) |
| Race, *n* (%) | White | 71 (73.2) | 38 (71.7) | 33 (75.0) |
|  | Black | 9 (9.3) | 5 (9.4) | 4 (9.1) |
|  | Asian | 3 (3.1) | 2 (3.8) | 1 (2.3) |
|  | Mixed | 5 (5.2) | 3 (5.7) | 2 (4.5) |
|  | Other | 9 (9.3) | 5 (9.4) | 4 (9.1) |
| Menopausal status, *n* (%) | Pre | 81 (83.5) | 48 (90.6) | 33 (75.0) |
|  | Post | 16 (16.5) | 5 (9.4) | 11 (25.0) |
| Stage, *n* (%) | I | 21 (21.6) | 8 (15.1) | 13 (29.5) |
|  | II | 60 (61.9) | 37 (69.8) | 23 (52.3) |
|  | III | 16 (16.5) | 8 (15.1) | 8 (18.2) |
| Pre-treatment node status, *n* (%) | Positive | 41 (42.3) | 23 (43.4) | 18 (40.9) |
|  | Negative | 56 (57.7) | 30 (56.6) | 26 (59.1) |
| Histologic type, *n* (%) | IDC | 90 (92.8) | 50 (94.3) | 40 (90.9) |
|  | ILC | 4 (4.1) | 2 (3.8) | 2 (4.5) |
|  | IDC and ILC | 2 (2.1) | 1 (1.9) | 1 (2.3) |
|  | Other | 1 (1.0) | 0 (0.0) | 1 (2.3) |
| Tumor grade, *n* (%) | 1 | 21 (21.6) | 12 (22.6) | 9 (20.5) |
|  | 3 | 76 (78.4) | 41 (77.4) | 35 (79.5) |
| TNBC, *n* (%) | Yes | 64 (66.0) | 36 (67.9) | 28 (63.6) |
|  | No | 33 (34.0) | 17 (32.1) | 16 (36.4) |
| Lymphovascular invasion, *n* (%) | Yes | 17 (17.5) | 12 (22.6) | 5 (11.4) |
|  | No | 80 (82.5) | 41 (77.4) | 39 (88.6) |
| TILs, *n* (%) | Absent/Scant | 58 (59.8) | 32 (60.4) | 26 (59.1) |
|  | Moderate/Marked | 38 (39.2) | 21 (39.6) | 17 (38.6) |
|  | Unknown | 1 (1.0) | 0 (0.0) | 1 (2.3) |
| pCR, *n* (%) | Yes | 22 (22.7) | 11 (20.8) | 11 (25.0) |
|  | No | 75 (77.3) | 42 (79.2) | 33 (75.0) |
| RCB, *n* (%) | 0/1 | 40 (41.2) | 20 (37.7) | 20 (45.5) |
|  | 2/3 | 57 (58.8) | 33 (62.3) | 24 (54.5) |

AC, doxorubicin-cyclophosphamide; SD, standard deviation; IDC or ILC, invasive ductal or lobular carcinoma; TNBC, triple negative breast cancer defined as ≤1% estrogen and progesterone receptor immunoreactivity; TILs, tumor infiltrating lymphocytes; pCR, pathologic complete response; RCB, residual cancer burden.

Among the 53 patients who received cisplatin, three miRNAs were associated with the primary outcome of achieving pCR while 10 miRNAs were associated with the secondary outcome of attaining minimal residual disease RCB score of 0/1 (both *p*<0.01 and FDR<0.25; **Table S1**). MiR-362-3p was the only miRNA associated with both outcomes (**Fig. 2A**). MiR-362-3p expression was 1.8-fold higher in patients who achieved a pCR and 1.7-fold higher in those attaining RCB 0/1 compared with non-responders (**Fig. 2B**). The association of miR-362-3p with pCR and RCB 0/1 remained significant in the fully adjusted multivariable models accounting for age at surgery, year of enrollment, clinical stage, tumor grade, TNBC status, and tumor-infiltrating lymphocytes (TILs; *p*=0.011 and *p*=0.008, respectively; **Table S2**). The multivariable logistic regression model used log2-transformed 2^-ΔCt^ values to demonstrate that higher miR-362-3p levels were significantly associated with increased odds of response (pCR: odds ratio (OR) 5.00 per 1-unit increase, 95% confidence interval [CI] 1.69-22.1; RCB 0/1: OR 4.63, 95% CI 1.72-17.7). No miRNA was associated with pCR or RCB 0/1 at *p*<0.01 and FDR<0.25 in the AC arm or among all 97 patients (**Table S1**). The exclusive association of miR-362-3p with clinical response in the cisplatin arm but not in the AC cohort suggests that this miRNA is a specific determinant of platinum sensitivity.

**Fig. 2.**
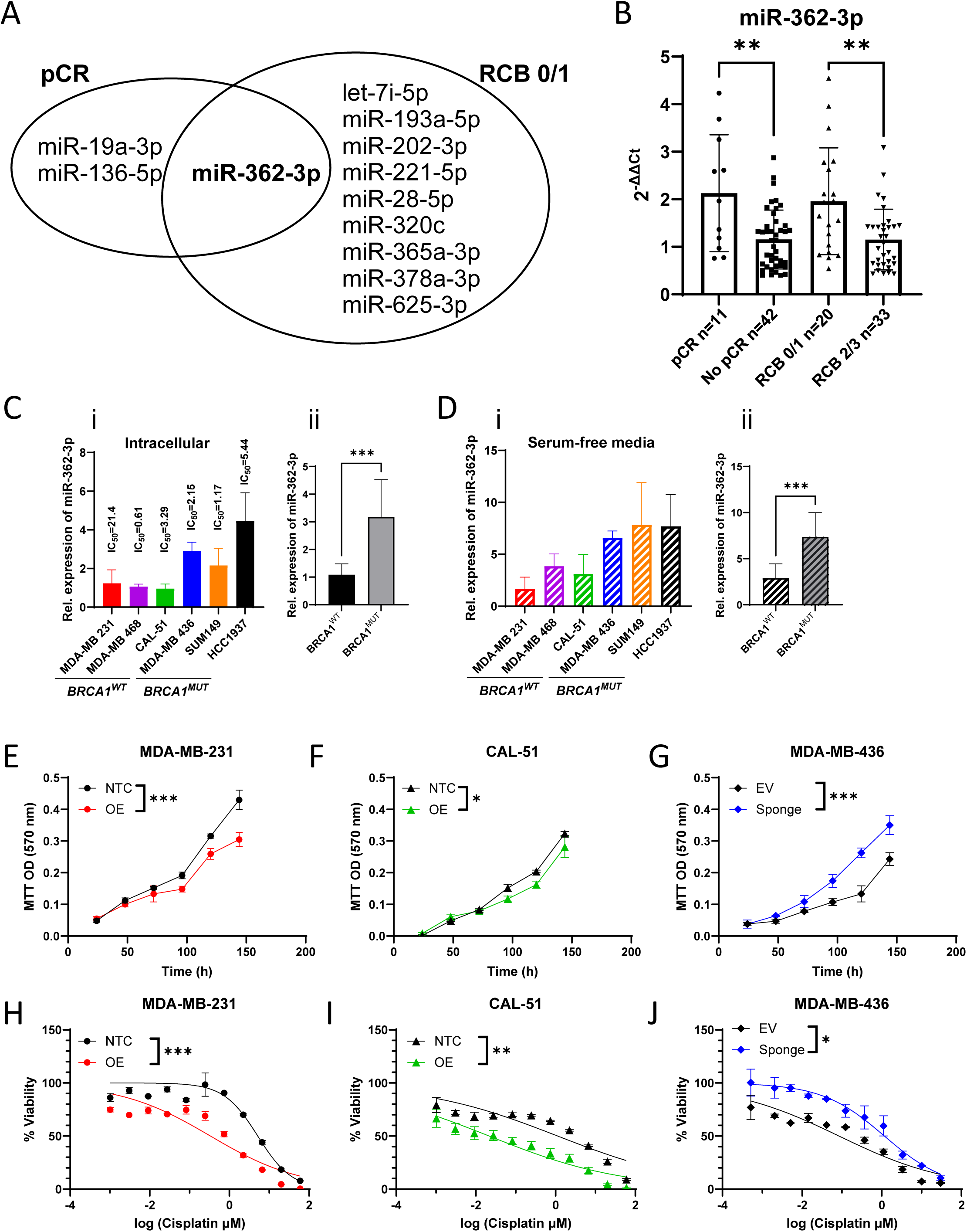
MiR-362-3p expression is enriched in *BRCA1^MUT^* TNBC and sensitizes cells to cisplatin. **A.** Discovery of plasma miRNAs associated with pathologic complete response (pCR) or minimal residual cancer burden (RCB) score of 0/1 in cisplatin-treated patients at *p*<0.01 and FDR<0.25; miR-362-3p was the only miRNA associated with both clinical outcomes. **B.** Relative plasma miR-362-3p levels stratified by pCR and RCB status. **C-D.** Intracellular (**C**) and extracellular (**D**) miR-362-3p expression in TNBC cell lines, stratified by *BRCA1*^MUT^ status. Data normalized to *U6* (cellular) or miR-16 (conditioned media). **E-G.** Effect of miR-362-3p overexpression (OE; **E**, **F**) or sponge-mediated inhibition (**G**) on cellular proliferation in indicated TNBC models compared to non-targeting controls (NTC) or empty vector (EV). **H-J.** Dose-response curves illustrating *IC*_50_ shirts following miR-362-3p OE (**H**, **I**) or inhibition (**J**) in response to cisplatin treatment. Data represent mean±SD in bar graphs and mean±SEM in dose response curves. *, ** and *** represent *p*<0.05, *p*<0.01, and *p*<0.001, respectively.

### Tumor miR-362-3p expression was not associated with cisplatin response

Small RNA sequencing was performed on baseline INFORM tumor biopsies; matching plasma miRNA profiles were successfully obtained for 79 of the 97 patients (**Fig. 1**). In this manuscript, we focus exclusively on tumor miR-362-3p expression; analyses of other tumor miRNAs are reported elsewhere. While miR-362-3p was expressed within the tumors, these levels did not correlate with their corresponding plasma concentrations (all *p*>0.05, **Fig. S1**). Tumor miR-362-3p was also not associated with pCR or RCB 0/1 in all patients and within each treatment arm (all *p*>0.05). Hence, the association between miR-362-3p and cisplatin sensitivity is specific to circulating miR-362-3p.

We then investigated miR-362-3p expression in TCGA. MiR-362-3p expression in breast tumors was two-fold higher compared to adjacent normal breast tissue (*p*<0.001; **Fig. S2A**). This finding remained significant even after accounting for age at surgery, year of recruitment, paired sampling, clinical stage, grade, and ER status (adj *p*<0.001). Within TNBC and ER+/PR+ subtypes, miR-362-3p expression in tumors remained higher than normal tissue (TNBC: 1.6-fold higher, *p*=0.07; ER+/PR+: 1.9-fold higher, *p*<0.001; **Fig. S2B**). MiR-362-3p expression remained significantly higher in ER+/PR+ tumors than normal tissue in the fully adjusted model (*adj.p*<0.001). When comparing between subtypes, miR-362-3p expression was 1.6-fold higher in TNBC than ER+/PR+ tumors (*p*<0.001 and *adj.p*<0.001; **Fig. S2B**). We explored whether *BRCA1/2^MUT^* or Single Base Substitution 3 (SBS3) status influence tumor miR-362-3p levels. Tumor miR-362-3p expression did not differ by *BRCA1/2^MUT^* (*p*=0.12) or SBS3 status (*p*=0.43). We did not analyze the association between tumor miR-362-3p and cisplatin response because no TCGA participant was treated with cisplatin ^26^. The correlation of plasma, but not intra-tumoral levels of miR-362-3p, with response suggests that circulating levels of miR-362-3p are a more sensitive indicator of the biological state that dictates platinum response.

### TBCRC 030 study and plasma miRNA

We sought to validate the INFORM plasma miR-362-3p observation in an independent cohort. TBCRC 030 (NCT01982448) was a Phase II study comparing neoadjuvant cisplatin to paclitaxel in TNBC patients ^2^. MiRNA screening was performed on 89 pretreatment plasma samples (**Fig. S3**). The clinical and tumor characteristics of these 89 TBCRC 030 patients were comparable to INFORM study—mostly stage II disease (74.2%), 34.8% had lymph node involvement, 96.6% were invasive ductal carcinoma (96.6%) and mostly grade 3 (93.3%). Germline *BRCA1/2* mutations were present in only 4 patients (4.5%). Among cisplatin treated patients, pCR and RCB 0/1 were achieved in 13.0% and 23.9% of the cases, respectively (**Table S3**). The association between plasma miR-362-3p and clinical response observed in INFORM was not recapitulated in the TBCRC 030 cohort (**Table S4**). This may be due to the differences in clinical response rates to cisplatin, which was 61.5% higher in the INFORM than in TBCRC 030 (pCR: 20.8% versus 13%; RCB 0/1: 37.7% versus 23.9%). Our findings suggest that circulating miR-362-3p may not be a stand-alone predictive biomarker for platinum sensitivity, but they allow for the possibility that increasing miR-362-3p will functionally improve outcomes of platinum treatments.

### MiR-362-3p enhances cisplatin sensitivity *in vitro*

Given the predominance of TNBC in the INFORM cohort and the higher miR-362-3p levels observed in TNBC tumors in TCGA, we performed mechanistic studies in TNBC cell lines to define the functional role of miR-362-3p in modulating cisplatin response. First we screened six cell lines—three *BRCA1^MUT^* (SUM149PT, HCC1937, and MDA-MB-436) and three *BRCA^WT^*(MDA-MB-468, MDA-MB-231, and CAL-51)—to assess cisplatin sensitivity (**Figs 2C,S4A**), baseline miR-362-3p expression (**Fig. 2C**), and whether tumor cells secrete miR-362-3p into serum-free conditioned media (**Fig. 2D**). *BRCA1^MUT^* cells exhibited higher intracellular miR-362-3p expression and more miR-362-3p were detected in serum-free conditioned media compared with *BRCA1^WT^*cells (both *p*<0.001; **Fig. 2Cii, 2Dii**).

Based on cisplatin sensitivity and baseline miR-362-3p expression, we generated two gain-of-function and one loss-of-function TNBC models. We overexpressed miR-362-3p in MDA-MB-231 and CAL-51 cells, and used a sponge to inhibit miR-362-3p in MDA-MB-436 cells (**Fig. S4B-D**). MiR-362-3p overexpression reduced cell growth (**Figs. 2E,2F**) whereas inhibiting miR-362-3p led to increased cell growth when compared to controls (**Fig. 2G**). MiR-362-3p overexpression also increased sensitivity to cisplatin (**Figs. 2H,I**) while inhibition of miR-362-3p reduced cisplatin sensitivity (**Fig. 2J**). Our *in vitro* data established that miR-362-3p is not a correlative bystander but a functional component of cisplatin sensitivity.

### MiR-362-3p enhances cisplatin response *in vivo*

We next validated our *in vitro* findings using cisplatin-treated xenograft models (**Fig. S5**). Consistent with the observed *in vitro* phenotypes, xenografts derived from MDA-MB-231 and CAL-51 cells overexpressing miR-362-3p exhibited significantly slower growth and prolonged survival in tumor-bearing mice following cisplatin treatment compared with control tumors (*p*=0.04 and *p*=0.002, respectively; **Figs. 3A,3B**,**S6A-D**). In contrast, miR-362-3p extinction in MDA-MB-436 cells resulted in accelerated tumor growth and shortened tumor-related survival following cisplatin treatment (*p*<0.001; **Figs. 3C,S6E-I**). These data show that miR-362-3p is an effective modifier of platinum efficacy, suggesting that mimicking its activity could be a viable strategy to enhance cisplatin sensitivity in TNBC.

**Fig. 3.**
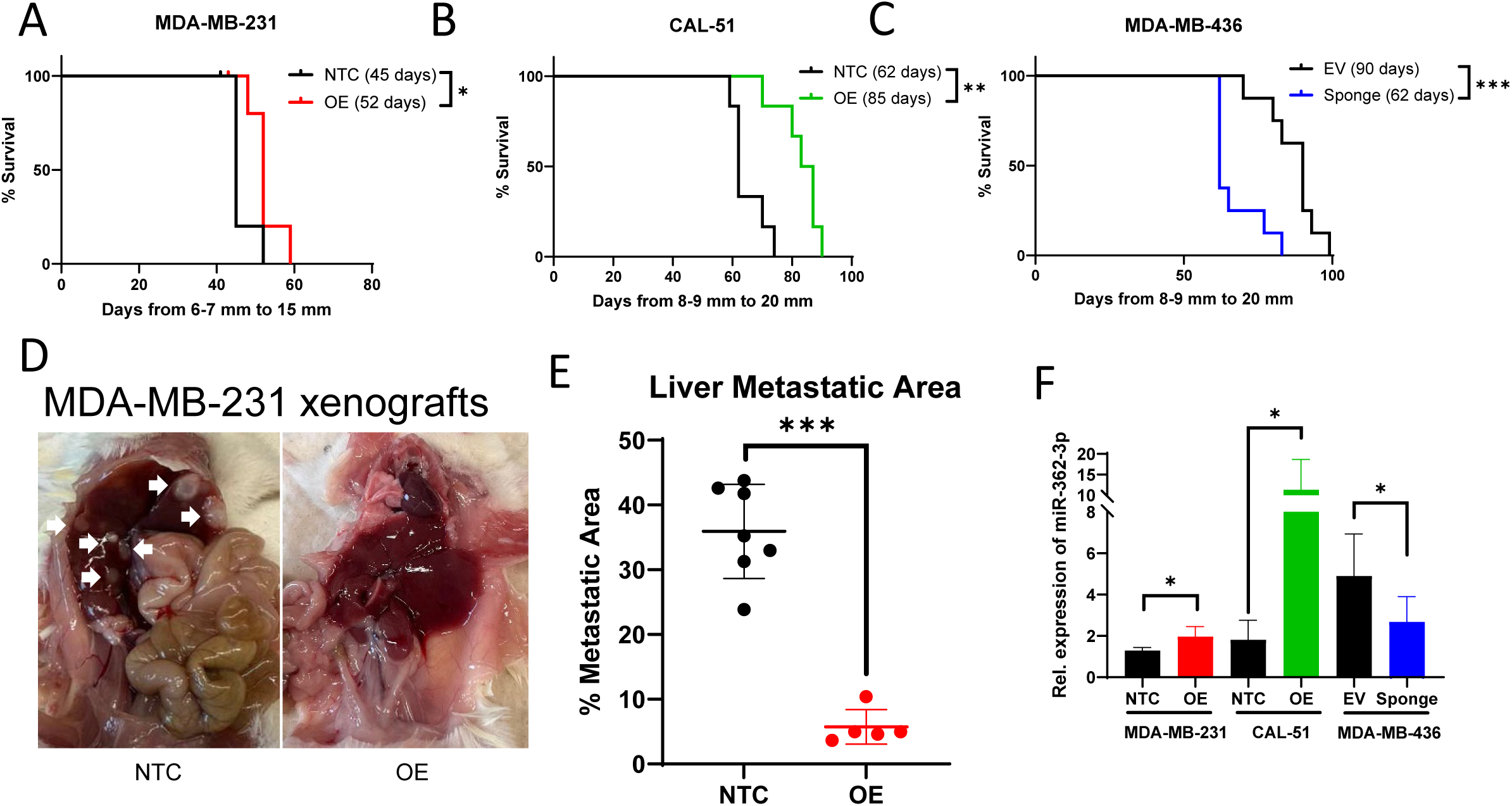
MiR-362-3p suppresses liver metastasis and sensitizes TNBC xenografts to cisplatin *in vivo*. **A-C.** Kaplan-Meier survival analysis of mice bearing MDA-MB-231 (**A**), CAL-51 (**B**), or MDA-MB-436 (**C**) tumors treated with cisplatin. Median survival was extended by miR-362-3p overexpression (OE) and shortened by miR-362-3p-inhibition compared to respective controls. **D.** Representative gross morphology of livers at necropsy; white arrows indicate macroscopic metastatic nodules in non-targeting controls (NTC) vs. miR-362-3p-OE. **E.** Quantitative of metastatic burden from H&E stained liver sections in MDA-MB-231 models. **F.** qRT-PCR validation of miR-362-3p levels in endpoint xenograft tumors, confirming sustained OE or inhibition throughout the study. Data represent mean±SD. *, ** and *** represent *p*<0.05, *p*<0.01, and *p*<0.001, respectively. Empty vector, EV.

### MDA-MB-231 cells overexpressing miR-362-3p exhibit reduced liver metastatic burden

Our orthotopically implanted MDA-MB-231 cells not only formed tumors, but also metastasized to liver and lung as has been observed before. Notably, liver tissues from mice bearing miR-362-3p-overexpressing MDA-MB-231 xenografts had markedly fewer macroscopic metastatic nodules, with no detectable liver metastases in a subset of animals upon necropsy (**Fig. 3D**). These gross observations were further confirmed by hematoxylin and eosin (H&E) staining (**Fig. S7**) and quantitative image analysis (*p*<0.001; **Fig. 3E**). In contrast, metastatic burden in the lung was unaffected by miR-362-3p status (*p*=0.20; **Fig. S8**). Analysis of tumors collected at experimental endpoints confirmed sustained modulation of miR-362-3p expression *in vivo* (**Fig. 3F**), supporting the stability of miRNA manipulation throughout the course of treatment. Our data demonstrate that miR-362-3p exerts organ-specific anti-metastatic effects, selectively compromising the ability of MDA-MB-231 cells to colonize the liver while leaving lung metastatic potential intact.

### *BCLAF1* is a direct target of miR-362-3p

To understand the mechanism underlying miR-362-3p-mediated enhancement of cisplatin sensitivity in TNBC, we first performed integrated analyses to identify candidate direct targets of miR-362-3p. Analysis of TCGA data identified 94 genes whose expression significantly correlated with miR-362-3p in both the full cohort of breast tumors (|ρ|>0.25, FDR<0.05) and the TNBC subset (|ρ|>0.25, *p*<0.01; **Table S5**). Of the 94 genes, only two genes—*BCLAF1* (BCL2-associated transcription factor 1) and *SSLC44A1* (choline transporter-like protein 1)—were predicted as direct targets by both TargetScanHuman ^27^ and miRDB ^28^ (**Table S5, S9A**). *BCLAF1* and *SLC44A1* mRNAs were negatively correlated with miR-362-3p in TCGA (**Figs. 4A,S9B-D**). *BCLAF1* is involved in DNA damage repair (DDR) ^29^, and its overexpression is linked to cisplatin resistance in lung cancer ^24^. *SLC44A1* is involved in cell proliferation and choline metabolism, and its inhibition promotes apoptosis in lung and colon cancer cells ^30,31^.

**Fig. 4.**
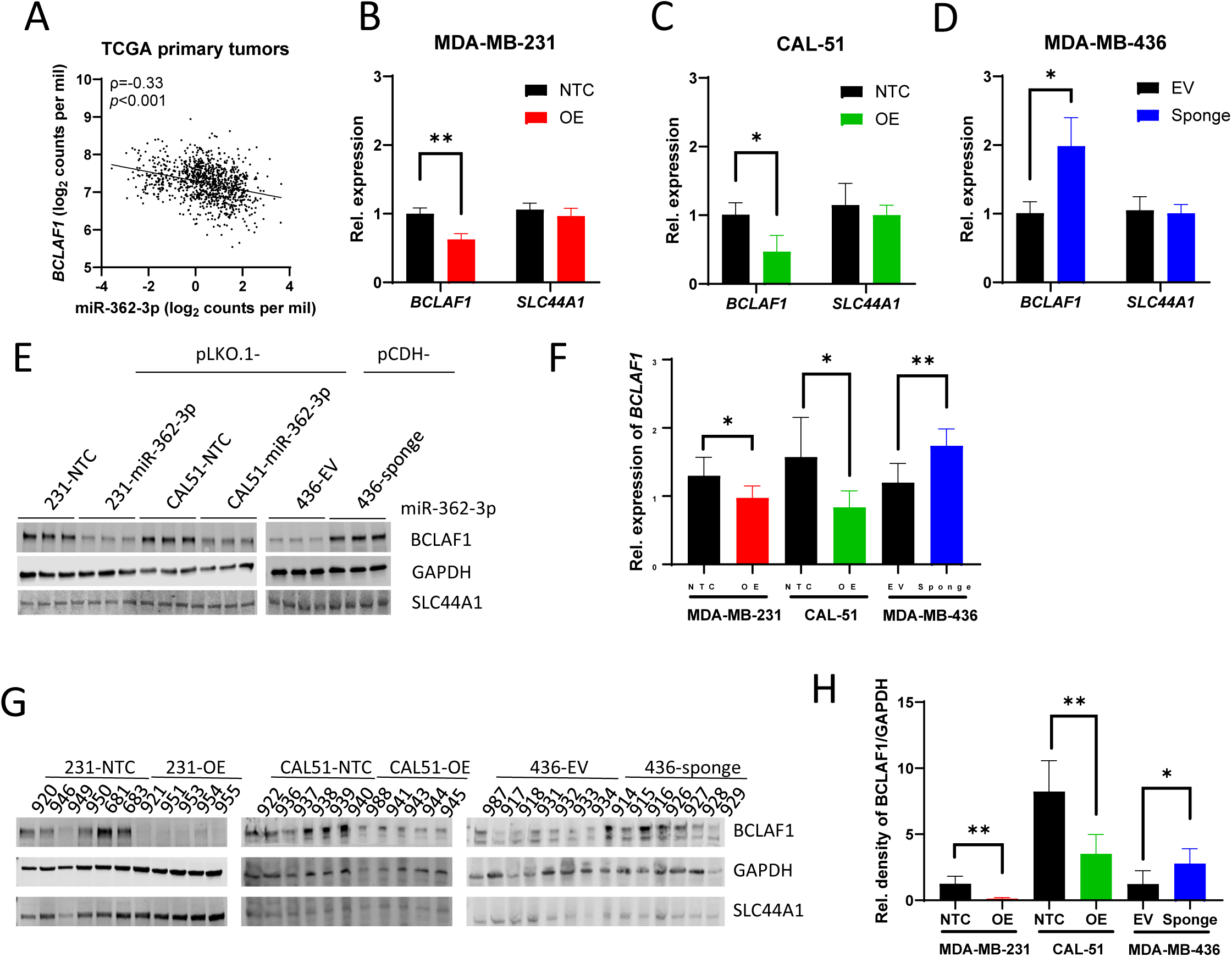
*BCLAF1* is a functional effector and direct target of miR-362-3p. **A.** Negative correlation between miR-362-3p and *BCLAF1* expression in TCGA breast cancer cohort. **B-D.** qRT-PCR analysis of *BCLAF1* and *SLC44A1* mRNA levels following miR-362-3p modulation *in vitro*. MiR-362-3p levels inversely suppressed *BCLAF1* mRNA, while SLC44A1 remains unaffected. **E.** Western blotting demonstrating reciprocal changes in BCLAF1 protein expression, but not SLC44A1, upon miR-362-3p overexpression (OE) or inhibition. **F-G.** Validation of miR-362-3p-mediated regulation of *BCLAF1* mRNA (**F**) and protein (**G**) in endpoint xenograft tumors. **H** The densitometric quantification of the 145 kDa BCLAF1 isoform (upper band). Data represent mean±SD. * and ** represent *p*<0.05 and *p*<0.01, respectively. Non-targeting controls, NTC; empty vector, EV.

Next, we assessed whether miR-362-3p directly regulates these candidate targets *in vitro*. Modulation of miR-362-3p expression resulted in robust and reciprocal changes in *BCLAF1* at both the mRNA and protein levels (**Figs. 4B-E**), confirming *BCLAF1* as a functional target of miR-362-3p. MiR-362-3p overexpression significantly attenuated *BCLAF1* expression, whereas miR-362-3p inhibition increased *BCLAF1* levels. In contrast, *SLC44A1* expression was not altered by miR-362-3p manipulation (**Figs. 4B-E**). The effect of miR-362-3p on *BCLAF1* persisted in tumors *in vivo* (**Figs. 4F-H**), confirming sustained target regulation over the course of treatment. The modulation of *BCLAF1* by miR-362-3p was consistent in tumors collected at experimental endpoints (**Figs. 4F-H**), confirming that the miR-362-3p/*BCLAF1* regulatory axis remains operative *in vivo*. By integrating bioinformatics target prediction with experimental validation across multiple models, we identify *BCLAF1* as a functional effector of miR-362-3p, and establish a potential regulatory link between the miRNA and a known mediator of DNA damage signaling ^24,29^.

### MiR-362-3p*/BCLAF1* axis regulates cisplatin efficacy

Given *BCLAF1* facilitates DNA damage signaling and repair, we assessed cisplatin-induced DNA damage by measuring γ-H2AX levels. As expected, cisplatin increased γ-H2AX *in vitro*, and this effect was significantly potentiated by miR-362-3p overexpression in MDA-MB-231 and CAL-51 cells (**Fig. 5A**). MiR-362-3p inhibition in MDA-MB-436 cells markedly attenuated cisplatin-induced γ-H2AX accumulation (**Fig. 5B**). Importantly, γ-H2AX levels were inversely correlated with BCLAF1 expression (**Figs. 5A,5B**).

**Fig. 5.**
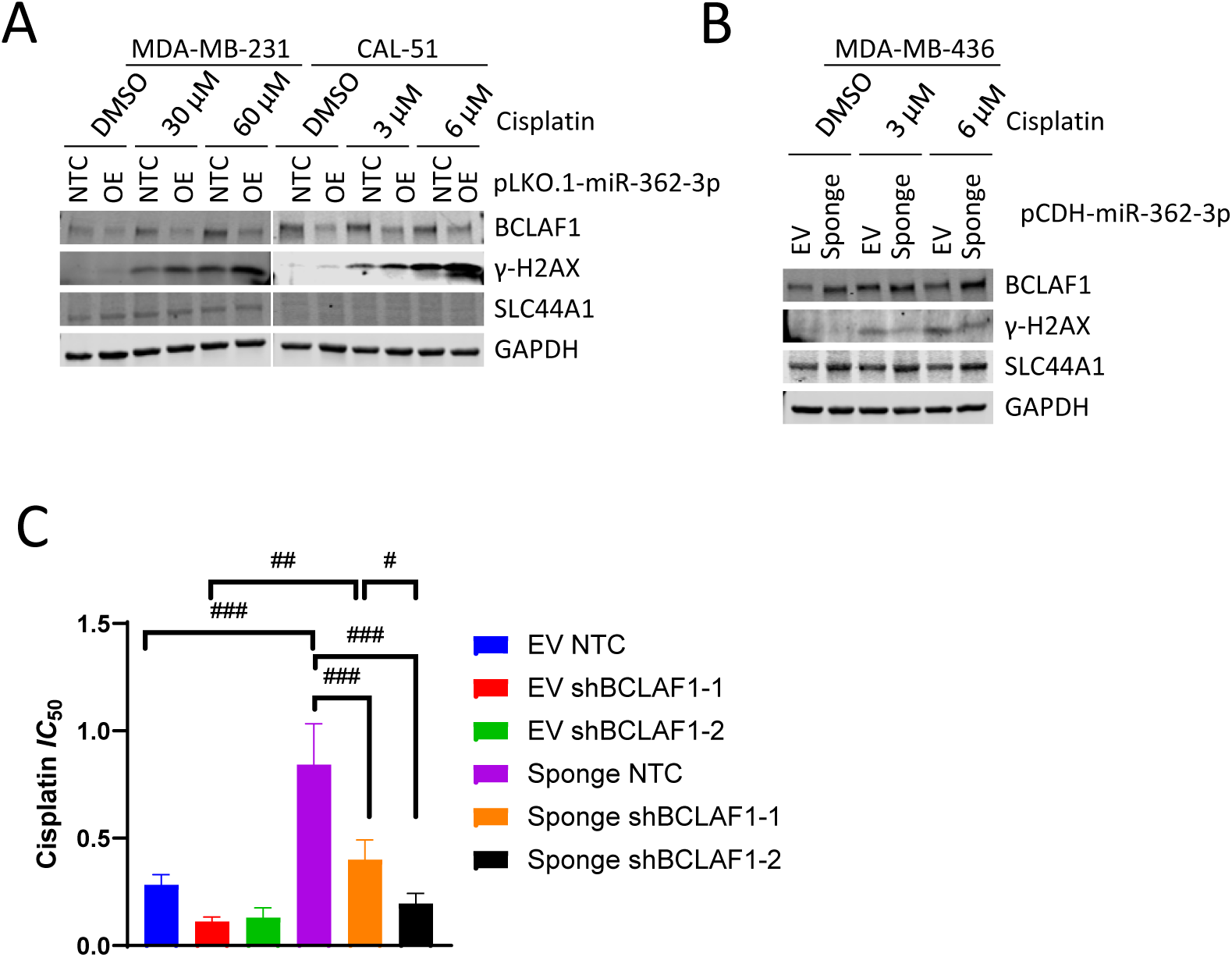
MiR-362-3p enhances cisplatin sensitivity by suppressing *BCLAF1*. **A.** Western blotting demonstrating that miR-362-3p overexpression (OE) suppresses *BCLAF1* and further potentiates cisplatin-induced γ-H2AX accumulation *in vitro*. **B.** Western blot showing that miR-362-3p inhibition in MDA-MB-436 cells attenuates γ-H2AX expression following cisplatin treatment. **C.** Functional rescue of cisplatin sensitivity in miR-362-3p-inhibited cells following *BCLAF1* knockdown using two independent shRNAs. Data represent mean±SD. ^#^, ^##^ and ^###^ represent *adj p*<0.05, *adj p*<0.01, and *adj p*<0.001, respectively. Non-targeting controls, NTC; empty vector, EV.

To definitively establish *BCLAF1* as a functional mediator of miR-362-3p in regulating cisplatin response, we performed epistasis experiments. *BCLAF1* was silenced using shRNAs in MDA-MB-436 cells (**Fig. S10A**). The cisplatin resistance induced by miR-362-3p inhibition was effectively reversed by the concomitant suppression of *BCLAF1* (**Figs. 4C,S10B**). Two-Way ANOVA revealed a significant interaction between miR-362-3p inhibition and *BCLAF1* status (*p*=0.002), confirming that the rescue observed with *BCLAF1* silencing was not merely additive, but rather a direct functional dependency within the miR-362-3p/*BCLAF1* axis. Our results support the miR-362-3p/*BCLAF1* axis as a regulator of cisplatin response in TNBC.

## Discussion

By bridging clinical trial data with functional validation, our study identified the miR-362-3p/*BCLAF1* axis as a previously unrecognized molecular determinant of platinum response in TNBC. MiR-362-3p functions as a sensitizer that dictates the cellular threshold for platinum-induced cell death by suppressing *BCLAF1*. Functionally, mir363-3p impairs the tumor’s capacity to repair platinum-induced DNA lesions. Furthermore, our findings reveal that miR-362-3p selectively suppresses liver metastasis, suggesting that this miRNA governs distinct programs of malignant progression and therapeutic vulnerability. Ultimately, the miR-363-3p/*BCLAF1* axis represents a targetable node that could be leveraged to overcome platinum resistance.

There was compartmental discordance between circulating and intratumoral miR-362-3p expression. While miR-362-3p was overexpressed in TNBC relative to normal breast, intratumoral levels did not correlate with paired plasma concentrations or clinical response. This lack of correlation between tumor and plasma samples is a recognized phenomenon in liquid biopsy research ^32^, often attributed to disparate tumor-intrinsic levels and complex systemic kinetics. In this setting, the plasma-based readout likely reflects an integrated signal of the tumor’s output, either active secretion or passive shedding during cellular turnover, in addition to contributions from the tumor microenvironment, systemic sources and the kinetics of microRNA clearing. The higher levels of miR-362-3p in tumors compared to adjacent normal tissue may represent compensatory response to oncogenic stress. While miR-362-3p typically functions as a tumor suppressor ^18^, its high levels may be insufficient to arrest tumor growth alone but may lower the threshold for apoptosis once cisplatin-induced DNA damage is introduced.

The functional significance of miR-362-3p lies in its suppression of *BCLAF1*. BCLAF1 is a DDR factor that is recruited to sites of DNA damage under the regulation of the BRCA1–BACH1 complex where it contributes to efficient homologous recombination repair ^29,33^. It is conceivable that miR-362-3p-mediated suppression of *BCLAF1* impairs the recruitment of the DNA repair machinery, thus enhancing platinum-induced cell death. This interaction may be particularly consequential in the context of *BRCA1/2* mutations. In HR-deficient *BRCA1/2^MUT^* cells, the functional loss of *BCLAF1* may serve as a second hit to the tumor’s repair capacity, preventing the resolution of stalled replication forks ^34,35^ and lowering the threshold for cell death. This “double-hit” model may explain why miR-362-3p was uniquely associated with a clinical response to cisplatin in the INFORM study but not in the TBCRC 030 cohort. In *BRCA1/2^MUT^*patients, the convergence of an inherited repair deficiency and a miRNA-induced “backup” repair failure ensures that platinum-induced stress leads to cell death rather than survival.

Our MDA-MB-231 xenograft studies revealed a striking divergence in how miR-362-3p/*BCLAF1* axis influences metastatic colonization. While the suppression of *BCLAF1* significantly reduced the formation and expansion of liver metastases, it had a negligible effect on lung metastases. This suggests that *BCLAF1*-mediated DNA repair is a prerequisite for TNBC cells to survive the hepatic microenvironment, and possibly allow them to survive the high-shear stress and exposure to reactive oxygen species in the portal circulation ^36,37^. When miR-362-3p is overexpressed, the impairment of *BCLAF1* function exacerbates genomic instability in cancer cells, which sensitizes to the anti-neoplastic effects of platinum and renders them unfit to survive the journey to, or colonize, the liver.

The miR-362-3p/*BCLAF1* axis represents a therapeutic vulnerability that could be leveraged to augment treatment of TNBC. Despite the clinical impact of KEYNOTE-522, approximately 36% of patients still have residual disease and remain at risk of recurrence ^4^, and for patients with metastatic TNBC, platinum agents are not curative ^38^ MiR-362-3p-based co-therapeutic could conceivably suppress *BCLAF1* and provide a mechanistic basis for overcoming platinum resistance. Furthermore, the synthetic lethal interaction between miR-362-3p-mediated *BCLAF1* inhibition and platinum-based chemotherapy holds promise beyond TNBC as it could potentially generalizable to other solid cancers where platinum remains the standard of care ^1^. Given that BCLAF1 complexes with BRCA1 to regulate in DDR pathways ^33^, the suppression of *BCLAF1* could confer a broad-spectrum vulnerability to other DNA-damaging modalities beyond platinum, including PARP inhibitors ^39^ and radiotherapy ^40^. In conclusion, our work defines the miR-362-3p/*BCLAF1* axis as a molecular determinant of platinum response and suggests that it may be exploited to overcome therapeutic resistance and improve outcomes for TNBC patients.

## Materials and Methods

### INFORM study

The INFORM trial was previously reported ^3^. Briefly, the trial enrolled 118 patients with a germline *BRCA1/2* mutation, a tumor size of ≥1 mm, N0-2, HER2-negative invasive breast cancer. Tumor estrogen receptor (ER) and progesterone receptor (PR) status were determined using immunohistochemistry. Baseline plasma and tumor biopsies were obtained when possible before initiating chemotherapy. Patients were randomly assigned 1:1 to either cisplatin 75 mg/m^2^ intravenously (IV) × 4 cycles or to AC (doxorubicin 60 mg/m^2^ IV and cyclophosphamide 600 mg/m^2^ IV) × 4 cycles every 2 (dose dense) or 3 weeks. After 4 treatment cycles, patients underwent mastectomy or lumpectomy, and sentinel lymph node biopsy or axillary dissection was performed.

### INFORM pathology review

The baseline tumor biopsy was assessed for histologic type and grade, lymphovascular invasion, and tumor infiltrating lymphocytes (TILs) using the International TILs Working Group criteria ^41^. Stromal TILs were scored as absent/scant for patients with < 25% stromal TILs or moderate/marked for patients with ≥ 25% stromal TILs ^42^.

Study endpoint tumors were assessed for residual cancer. Residual disease in the breast was assessed for overall cancer cellularity (%), tumor bed area, and percentage of the cancer that was *in situ* disease. Residual disease in axillary nodes was assessed for number of involved nodes and diameter of the largest metastasis. pCR in the breast was defined as the absence of residual invasive disease with or without ductal carcinoma *in situ* (DCIS). Patients with pCR in the breast and negative pretreatment sentinel lymph node biopsy were considered to have achieved pCR. The RCB score was determined using the online tool ^43^. RCB score of 0 is synonymous to achieving pCR. RCB score of 1, 2, and 3 corresponds to minimal, moderate, and extensive residual disease. The primary outcome was achieving pCR (i.e., pCR versus no pCR). The secondary outcome was assessing the proportion of patients with RCB 0/1 versus RCB 2/3.

### INFORM plasma miRNA screening

MiRNA screening was performed by the Detection Unit, Precision RNA Medicine Core at Beth Israel Deaconess Medical Center, Boston, MA. Baseline single-spun plasma samples were thawed on ice and spun twice. Total RNA was isolated from 200 µL of plasma using the Maxwell RSC miRNA Plasma and Serum Kit (Promega, Madison, WI, USA). During RNA extraction, 10 µL of ID3EAL™ Cancer Knowledge Panel spike-ins (Mirxes, Singapore) were added to each sample to monitor the efficiency of the RNA isolation and to normalize for technical variations during RNA isolation ^44^. Total RNA was eluted in 60 µl of RNase-free water.

The ID3EAL™ Cancer Knowledge Panel assay is a quantitative PCR (qPCR)-based method that quantifies 352 miRNAs in each sample on a 384-well plate. Briefly, cDNA synthesis was performed using 20 uL of isolated RNA at 42 C for 30 min, followed by heat-inactivation at 95 C for 5 min on the SimpliAmp^TM^ Thermal Cycler (Thermo Fisher Scientific, Waltham, MA, USA). Next, 12 uL of cDNA was augmented as recommended by the manufacturer using the cycling protocol of 95 C for 5 min, 40 C for 5 min, and 8 cycles of 95 C for 10s and 60 C for 40 s on the SimpliAmp^TM^ Thermal Cycler. Lastly, qPCR was performed on 25 uL of augmented cDNA on two Quant Studio 5 Real-Time PCR Systems (Thermo Fisher Scientific) over 13 days. The cycling conditions were 95 C for 10 min, followed by 40 cycles of 95 C for 10 s and 60 C for 30 s.

Per standard qPCR methodology, data points with raw Ct values of <8 or ≥33 were excluded. Samples with a RNA Spike-In control Ct value that exceeded 3 standard deviation (sd) from the mean of 9.9 were considered as failed assays and excluded. The Ct value of each miRNA was first normalized to the average scaling factor of interplate calibrators, followed by the scaling factor of RNA spike-ins for each plate. In order to further perform global geometric mean normalization ^45^, 101 miRNAs expressed in <10% of the samples were excluded, and missing data points were replaced using the mean ± 3 sd of the respective miRNA. ComBat (sva: Surrogate Variable Analysis, R package version 3.38.0) ^46^ was used to reduce batch effects as the samples were assayed across multiple days. Fold changes in miRNA expression were calculated using the 2^^-ΔΔCt^ method.

### INFORM Paired-tumor smallRNASeq

Total RNA was isolated from baseline tumor biopsies using the mirVana miRNA Isolation Kit (ThermoFisher Scientific) adhering to the manufacturer’s instructions. Extracted RNA was quantified (Qubit 4.0 Fluorometer, Life Technologies, Carlsbad, CA, USA), RNA integrity was assessed (TapeStation 4200, Agilent Technologies, Palo Alto, CA, USA), and ribosomal RNA depletion was performed (FastSelect Human/Mouse/Rat probe, QIAGEN, Germantown, MD, USA).

Small RNA sequencing libraries were prepared using Illumina TruSeq Small RNA library Prep Kit (Illumina, San Diego, CA, USA). In brief, Illumina 3’ and 5’ adapters were added to RNA molecules with a 5’phosphate and a 3’-hydroxyl group sequentially. Single-stranded cDNA was generated via reverse transcription and subsequently amplified using a common primer and an index-containing primer. The desired sequencing library was obtained via polyacrylamide gel electrophoresis of the amplified cDNA and excising a band ∼145–160 bp and concentrated via ethanol precipitation. The sequencing library was validated on the TapeStation 4200 (Agilent Technologies), and quantified by with a Qubit 4.0 Fluorometer (Invitrogen) and by qPCR (KAPA Biosystems, Wilmington, MA, USA).

Sequencing libraries were pooled, clustered, and loaded onto the Illumina sequencer according to the manufacturer’s instructions. Sequencing was performed using paired-end configuration. Image analysis and base calling were carried out using the Illumina HiSeq Control Software. Raw sequencing data (.bcl files) were converted to FASTQ format and de-multiplexed using bcl2fastq version 2.17 (Illumina). One mismatch was allowed for index sequence identification.

Raw small RNA-Seq reads were quality-checked using FastQC (version 0.11.8) ^47^ and preprocessed with Cutadapt (version 2.6) for adapter removal ^48^. Preprocessed reads were quantified using Manatee (version 1.0) ^49^, which maps reads to the human genome (GRCh38, Ensembl version 98) and transcriptome using Bowtie (version 1.0.1) ^50^before assigning reads to annotated regions. MiRNA sequences were retrieved from miRBase version 22 ^51^. Raw miRNA counts were transformed to log_2_ counts per million (CPM) and normalized using the voom function form the limma R package (version 3.58.1) ^52^. Differential expression was performed using limma ^53^.

### The Cancer Genome Atlas (TCGA) invasive breast cancer miRNA dataset

TCGA clinical, miRNA, and mRNA data were downloaded from Genomic Data Commons (GDC; release 42.0). MiRNA and mRNA data were mapped to Genome Build GRCh38. Of the 1207 available miRNA data files, 44 were excluded: 13 male samples, two DCIS, one did not have pathology review of the tumor, 12 had prior malignancy, and 16 had cancer therapy for a previous malignancy or neoadjuvant therapy, and 16 were technical or biological replicates. For samples with technical or biological replicates; the mean raw count for each miRNA was calculated to represent that sample, leading to the additional exclusion of 16 files. The remaining 1147 miRNA data consisted of 1038 primary tumors, 102 histologically normal-adjacent breast tissue, and 7 metastatic breast tumors from 1040 women. There were 100 tumor-normal pairs. Previously published tumor grade, germline *BRCA1/2* status, and single base substitution (SBS) signature 3 were retrieved ^54,55^. Of the 1038 primary tumors with miRNA data, 1036 had corresponding mRNA data.

### TBCRC 030 study

The primary objective of TBCRC 030 was to detect an association of HRD with RCB 0/1 to neoadjuvant cisplatin or paclitaxel ^2^. The trial concluded that the rate of RCB 0/1 did not differ between the treatment arms. While patients with a known germline *BRCA1/2^mut^* mutation at baseline were ineligible, *BRCA* status was later ascertained through commercial testing or via the HRD assay. Patients were randomized to receive either cisplatin 75 mg/m^2^ every 3 weeks x 4 cycles or weekly paclitaxel 80 mg/m^2^ for 12 weeks. After the completion of neoadjuvant therapy, patients underwent surgery. Patients with inadequate clinical response after 12 weeks, clinically or radiologically determined, “crossed over” to receive alternative provider-selected neoadjuvant therapy. Baseline plasma (*n*=110) and tumor biopsies were obtained when possible before initiating chemotherapy. Baseline tumor biopsy was assessed for histologic type and grade. Study endpoint tumors were assessed using the RCB score. Patients who crossed over after the 12 weeks of neoadjuvant therapy were considered non-responders and assigned as RCB 2/3. TBCRC 030 plasma samples were screened for miRNAs using the ID3EAL™ Cancer Knowledge Panel workflow as a validation cohort.

### TNBC cell lines

MDA-MB-231, MDA-MB-436, MDA-MB-468, and HCC1937 were acquired from the American Type Culture Collection (ATCC; Manassas, VA), CA51 (CVCL_1110) was obtained from Leibniz Institute DSMZ-German Collection of Microorganisms and Cell Cultures GmbH (Braunschweig, Germany) and SUM149 which was a gift from Dr. Christina Gewinner, Division of Signal Transduction, BIDMC. All cell lines were grown according to the providers’ instructions in the following manner: MDA-MB-231, MDA-MB-436, MDA-MB-468, and CAL-51 were propagated in Gibco™ DMEM, High Glucose (ThermoFisher Scientific) with 10% fetal bovine serum (FBS; GeminiBio, West Sacramento, CA); HCC1937 was propagated in RPMI 1640 (ThermoFisher Scientific) with 10% FBS and 2 mM L-glutamine; and SUM149 cells were grown in Ham’s F-12 medium (ThermoFisher Scientific) supplemented with 5% heat-inactivated FBS, insulin (5 μg/ml), and hydrocortisone (2 μg/ml). All cell lines were verified with short, repeating DNA (STR) profiling, tested for absence of mycoplasma and maintained in a humidified 5% CO_2_ incubator at 37°C.

### Plasmid transfection

MiR-362-3p overexpression plasmid pLKO.1-U6-PGK-GFP-p2a-puro-hsa-miR-362-3p, and knockdown plasmid pCDH-CMV-MCS-EF1a-mCherry-puro-miR-362-3p-sponges were commercially purchased (Paivibio, Wuhan, China). Each lentiviral plasmid was co-transfected with plasmids encoding Δ8.9 and VSVG into HEK293T packaging cells using jetPRIME® DNA and/or siRNA transfection reagent (Polyplus®, Illkirch, France). Viral supernatant was collected 48 hours post-transfection, filtered (0.22-nm pore size), and added to breast cancer cells in the presence of 8 mg/mL polybrene (Sigma-Aldrich, St Louis, MO) and stable cells were selected with puromycin (Sigma-Aldrich). Two short hairpin RNA (shRNA) plasmids targeting *BCLAF1* (TRCN0000336704, and TRCN0000020628), as well as non-targeting control shRNAs, cloned into the pLKO.1 lentiviral vector, were obtained from Sigma-Aldrich. *BCLAF1* shRNAs were introduced into miR-362-3p-knockdown MDA-MB-436 cells, and knockdown efficiency was confirmed by Western blotting prior to downstream assays.

### MTT assay for cell viability and proliferation

The MTT staining method (TargetMol Chemicals, Inc, Boston, MA) was used for cell viability and proliferation. Cells (1 to 5 × 10^3^/well) were plated in a 96-well plate and allowed to settle overnight, followed by treatment with DMEM/RPMI-1640/F-12 medium supplemented without or with various concentrations of Cisplatin (Sigma-Aldrich). The parental control or transfected cells were incubated at 37 °C and 5% CO_2_. After 24, 48, 72, 96, or 144-hour treatment, 10 μl of 5 mg/mL MTT reagent was added to the medium and cells were incubated for a further 4 h. The medium was then drained and cells were treated with 100 μL dimethyl sulfoxide for 15 min. The plates were then analyzed using a SpectraMax M5 Microplate Reader (Molecular Devices, LLC, San Jose, CA). A dose-response curve was generated by plotting the percentage of viability against cisplatin concentration and determine the IC50 values using nonlinear regression analysis in Graphpad Prism.

### Quantitative real time PCR (qRT-PCR)

For detection of miR-362-3p secreted into media, 10^^4^ cells were seeded in 60-mm cell culture dish and allowed to settle overnight, followed by treatment with serum-free DMEM for 24h. Serum-free media and breast cancer cell lines were collected for RNA isolation using miRNeasy Kit for miRNA Purification/RNeasy Mini Kit (Qiagen) according to the manufacturer’s instructions. RNA for target genes were subjected to cDNA synthesis with random hexamer and oligo(dT)_20_VN primers using HiScript III 1st Strand cDNA Synthesis Kit (+gDNA wiper) (Vazyme Biotech, Nanjing, China). MiRNA was subjected to cDNA synthesis with stem-loop primers using miRNA 1st Strand cDNA Synthesis Kit (Vazyme). qRT-PCR was performed on a 7500 HT Fast RT-PCR System (ThermoFisher Scientific) using a Taq Pro Universal SYBR qPCR Master Mix kit (Vazyme). The SYBR primer pair sequences were available in Table S6. Expression fold change was calculated using 2^-ΔΔCt^ method by first normalizing to *RPL13A* (for mRNA), *U6* (for miRNA), or miR-16 (for serum-free miRNA) ^56^ then further normalized to the average of the untreated controls.

### Immunoblotting

Cells were lysed in RIPA buffer supplemented with protease inhibitors (Roche). Protein concentrations were determined using the Bradford Protein Assay Kit (Pierce). Equal amounts of protein were resolved by SDS–PAGE (Bio-Rad) and transferred to PVDF membranes. Immunoblotting was performed following the manufacturer’s protocol using the LI-COR detection system. Membranes were incubated with the following primary antibodies: GAPDH (14C10) rabbit monoclonal antibody (Cell Signaling Technology, #2118, RRID:AB_561053), BCLAF1 polyclonal antibody (Proteintech Group, Inc, Rosemont, IL, #26809-1-AP, RRID:AB_2880642), phospho-Histone H2A.X (Ser139) (20E3) rabbit monoclonal antibody (Cell Signaling Technology, #9718, RRID:AB_2118009), and SLC44A1 polyclonal antibody (Proteintech Group, Inc, #33135-1-AP, RRID: AB_10696180). Appropriate IRDye-conjugated secondary antibodies were used for detection, and signals were visualized and quantified using the LI-COR imaging system.

### Animal experiments

All animal experiments were conducted in accordance with Institutional Animal Care and Use Committee-approved protocols at Beth Israel Deaconess Medical Center (052-2020-23). Female NSG mice (6–8 weeks of age) were used for xenograft studies. MDA-MB-231, MDA-MB-436, and CAL-51 breast cancer cells were cultured under standard conditions and harvested during logarithmic growth. Cells were washed twice with sterile phosphate-buffered saline (PBS) and resuspended in serum-free medium at a concentration of 1 × 10 cells per 100 μL. For miRNA manipulation, MDA-MB-231 and CAL-51 cells stably overexpressing miR-362-3p, and MDA-MB-436 cells with stable miR-362-3p knockdown, were generated by lentiviral transduction prior to implantation. Corresponding control cells were generated using empty vector or non-targeting constructs.

Each mouse was subcutaneously injected with 1 × 10 cells in 100 μL of serum-free medium into the flank. Tumor volume and body weight were measured twice weekly. Tumor volume was calculated using the formula: V = (length × width²) / 2. When tumors became palpable, mice were randomized into treatment groups. Cisplatin was administered to all mice by intraperitoneal injection once per week, using cell line-specific dosing regimens (Fig. S4). Dose schedules were selected based on preliminary tolerability observations and known differences in cisplatin sensitivity among the respective cell lines, with the goal of minimizing systemic toxicity while maintaining antitumor efficacy. Mice were monitored throughout the study for signs of treatment-related toxicity, including body weight loss, reduced activity, ruffled fur, and hunched posture. Mice exhibiting greater than 15% body weight loss or severe signs of distress were euthanized according to humane endpoint criteria.

Tumor measurements and endpoint assessments were performed in a blinded manner. At the experimental endpoint or upon reaching predefined humane criteria, mice were euthanized. Necropsy was performed to assess tumor metastatic burden. Liver and lung tissues were harvested, fixed in formalin, and embedded in paraffin for H&E staining. Tumors were excised and processed for protein and RNA extraction. Expression of miR-362-3p and *BCLAF1* was analyzed by quantitative RT-PCR and Western blotting.

### Metastatic burden evaluation in liver and lungs

Liver metastasis were morphologically distinct on H&E slides. Liver H&E slides (*n*=12) were digitized at 20× using the PhenoImager HT (Akoya Biosciences, Marlborough, MA). Metastatic burden was quantified using pixel classification in QuPath (version 0.6.0) ^57^. The first pixel classifier defined total tissue area using mostly default settings except: “Very low” resolution, Average channels, Smoothing sigma = 3, Threshold = 200, and Below threshold = “Tissue Area”. A second pixel classifier was used to define liver and metastatic areas using two training images, “Moderate” resolution, Region = “Any annotation bounds (fast)”, and default settings for the remainder. The two training images were created by compiling regions of interest (ROIs): one training image for metastatic area which comprised of 16 ROIs sampled across 12 images; and one training image for liver area, comprising of 12 ROIs total sampled across 12 images. Both classifiers were applied on all the images, and liver metastatic area was calculated as a percentage of total tissue area.

Lung metastases were not morphologically distinct on H&E slides. Therefore we performed Ku80 immunohistochemistry to detect metastases using the VECTASTAIN Elite ABC Universal PLUS Kit, Peroxidase (Vector Laboratories, Inc, Newark, CA). FFPE slides were deparaffinized with Histo-Clear (National Diagnostics, Thermo Fisher Scientific) and rehydrated through decreasing concentrations of ethanol to water. For heat-induced antigen retrieval, sections were heated in a steamer while submerged in Antigen Unmasking Solution (citric acid based) pH 6.0 for 15 min and cooled to room temperature for 45 min. Next, slides were rinsed in 1X PBS three times, for 5 min each time, and blocked with BLOXALL for 10 min. Slides were rinsed in 1X PBS three times, for 5 min each time, and blocked in 10% goat serum for 30 min and washed three times in 1X PBS once more. Afterwards, slides were incubated with rabbit monoclonal anti-human Ku80 clone C48E7 antibody (Cell Signaling Technology, Cat. #2180, RRID: AB_2218736; 1:1000 dilution) overnight at 4C. The next day, the slides were washed with 0.05% PBST three times, for 5 min each time, incubated with biotinylated goat anti-rabbit IgG (Vector Laboratories, #BP-9100-50, RRID:AB_3665891) for 30 min, once more washed with 0.05% PBST three times, and incubated with VECTASTAIN Elite ABC Reagent for 30 min. After, the slides were washed 3 times with 1X PBS, for 5 min each time and detection of antibodies was achieved by incubating slides with ImmPACT DAB EqV Substrate for 30 sec. Slides were immediately rinsed with MilliQ water and washed two more times with MilliQ water for 5 min each time. Slides were counterstained with Hematoxylin Solution (Mayer’s, Modified) for 30 sec, rinsed in 1X PBS for 5 min, and washed in 1X PBS for 5 min. Slides were dehydrated through increasing concentrations of ethanol to Histoclear and mounted using Paramount (Thermo Fisher Scientific).

Lung IHC slides (*n*=12) were digitized at 20× using the PhenoImager HT (Akoya Biosciences) and analyzed using QuPath using default values for most parameters except: hematoxylin threshold was set at 0.05 and nucleus mean optical density was set at 0.15 or 0.2 depending on the staining batch. Lung metastatic burden was calculated as a percentage of total cells detected.

### Statistical analysis

Fold changes in miRNA expression were calculated using the 2^^-ΔΔCt^ method. Binary logistic regression assessed the associations between plasma miRNA expression and pCR or RCB 0/1 among all patients and stratified by treatment arm. Because lower ΔCt values indicate higher miRNA abundance, we used log2(2^^−ΔCt^) in logistic regression models to make coefficients intuitively interpretable, such that a higher β corresponds to higher miRNA expression and increased odds of response. The minimally adjusted multivariable model accounted for age at surgery and year of enrollment while the fully adjusted multivariable model further incorporated for clinical stage, tumor grade, TNBC status, and tumor-infiltrating lymphocytes (TILs).

Tumor miRNA and mRNA raw counts from INFORM and TCGA were converted to log2 counts per million (CPM) values and normalized using voom (limma, R package version 3.58.1) ^52^. Differential expression was performed using limma ^53^. Correlation was performed using Spearman’s ρ. For TCGA, we compared miR-362-3p expression between primary tumors and normal breast tissue, and in a multivariable analysis controlling for age at surgery, year of recruitment, paired sampling, clinical stage, grade, and ER status. We also compared within subtype (TNBC: tumor *n*=112 versus normal *n*=9; ER+/PR+: tumor *n*=642 versus normal *n*=60) and between subtype (TNBC versus ER+/PR+ tumors) using unadjusted and fully adjusted models (age, year, pairing, stage, and grade). Additional analyses in TCGA tumors determined differential miRNA expression associated with germline *BRCA1/2^mut^*(*n*=117 versus non-*BRCA1/2 n*=921) and Single Base Substitution 3 (SBS) signature (yes *n*=128 versus no *n*=212). Multiple hypothesis correction were done by adjusting for false discovery rate (FDR) using the Benjamini & Hochberg ^58^. If statistical significance of FDR<0.05 was not achieved, meaningful findings were reported. Clinical plasma and tumor miRNA/mRNA analyses were conducted using R version 4.3.3. Statistical analysis for *in vitro* and *in vivo* studies were conducted using Graphpad Prism version 10.6.0 using Student’s *t* and Log-rank tests. For epistasis experiments, data were analyzed using Two-Way ANOVA to evaluate the interaction between miR-362-3p and *BCLAF1* modulation, followed by Tukey’s multiple comparisons test. Figures and graphs were plotted using Graphpad Prism and BioRender.

## Data Availability

All data produced in the present work are contained in the manuscript.

## Funding and Acknowledgements

This work was supported by NIH R35CA232105 (FJS), R01CA235740 (FJS), R01CA226259 (ALK), R01CA205420 (ALK), the Ludwig Center at Harvard (GMW), AGA/Jenzabar research fund (GMW), and Breast Cancer Research Foundation (NMT, GMW, JEG & SJS). Mirxes provided the ID3EAL™ Cancer miRNA Knowledge Panels. The Detection Unit, Precision RNA Medicine Core at BIDMC (RRID:SCR_024819), performed the ID3EAL™ assays. BIDMC Spatial Technologies Unit digitized the slides (RRID:SCR_024905) and the Histology Core provided histology services (RRID:SCR_009669). The authors thank the administrative staff for compiling the INFORM and TBCRC 030 trial data.

## Author Contributions

Conceived and designed the studies: YJH, GMW, FJS, ZL, SK. Clinical trial: NMT, JEG, SJS, ELM. Animal and molecular work: ZL, SK, LW, XL, BRS, EKA, ESM. Histology and image analysis: ZL, EKA, YJH. MiRNA assays and analysis: CW, MU, CSB, YJH, JKC, HC, LZ, PM. Tumor smallRNASeq and analysis: DK, NK, ISV, YJH. Data interpretation: YJH, GMW, FJS, ALK, ZL, FB, PM, JKC. All authors contributed to the writing and reviewing of the manuscript.

---

**Table S1.** The associations of 352 miRNAs with pCR **(A)** or RCB 0/1 **(B)** among all INFORM subjects, and when stratified by treatment.

**Table S2.** Multivariable analysis of miR-362-3p associations with pathologic complete response (pCR; **A**) and residual cancer burden (RCB) score of 0 or 1 **(B)** among 53 cisplatin-treated patients.

**Table S3.** Demographic and tumor characteristics of the TBCRC 030 participants, stratified by treatment.

**Table S4.** The associations of 352 miRNAs with pCR **(A)** or RCB 0/1 **(B)** among all TBCRC 030 subjects, and when stratified by treatment.

**Table S5.** Integrative analysis of TCGA and predictive databases to identify miR-362-3p candidate direct target genes.

**Table S6.** Primer sequences.

