## Supplementary material for "The miR-362-3p/*BCLAF1* axis regulates cisplatin sensitivity and metastatic progression in triple-negative breast cancer": Fig. S

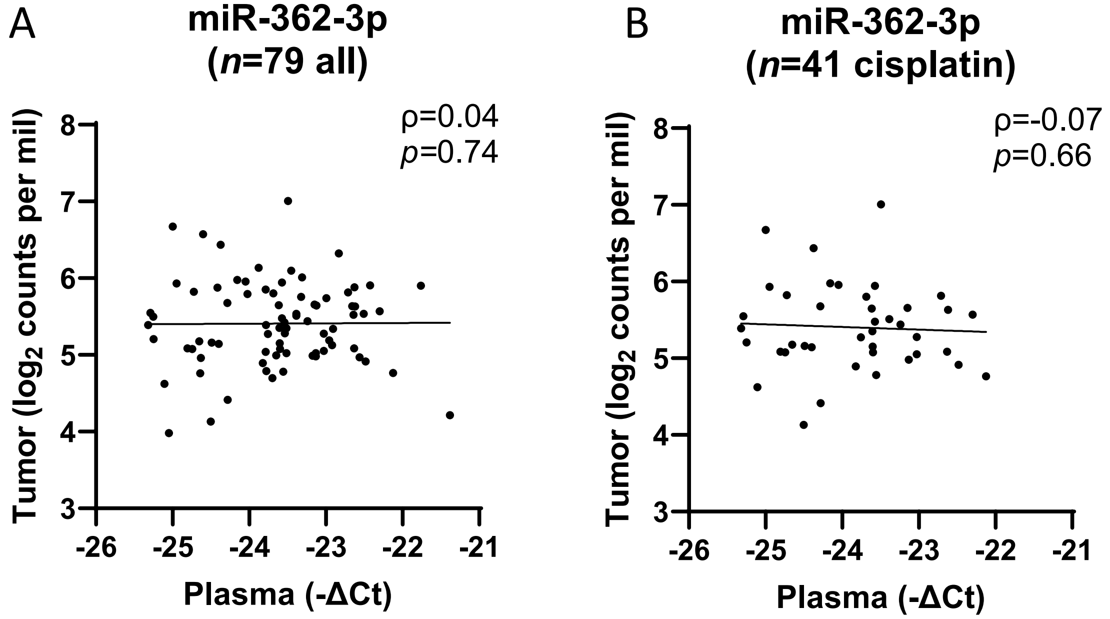


**Fig. S1. Correlation analysis of tumor and plasma miR-362-3p levels in INFORM.** No significant correlation was observed between the two compartments all 79 subjects (**A**) or in the subset of 41 patients treated with cisplatin (**B**).

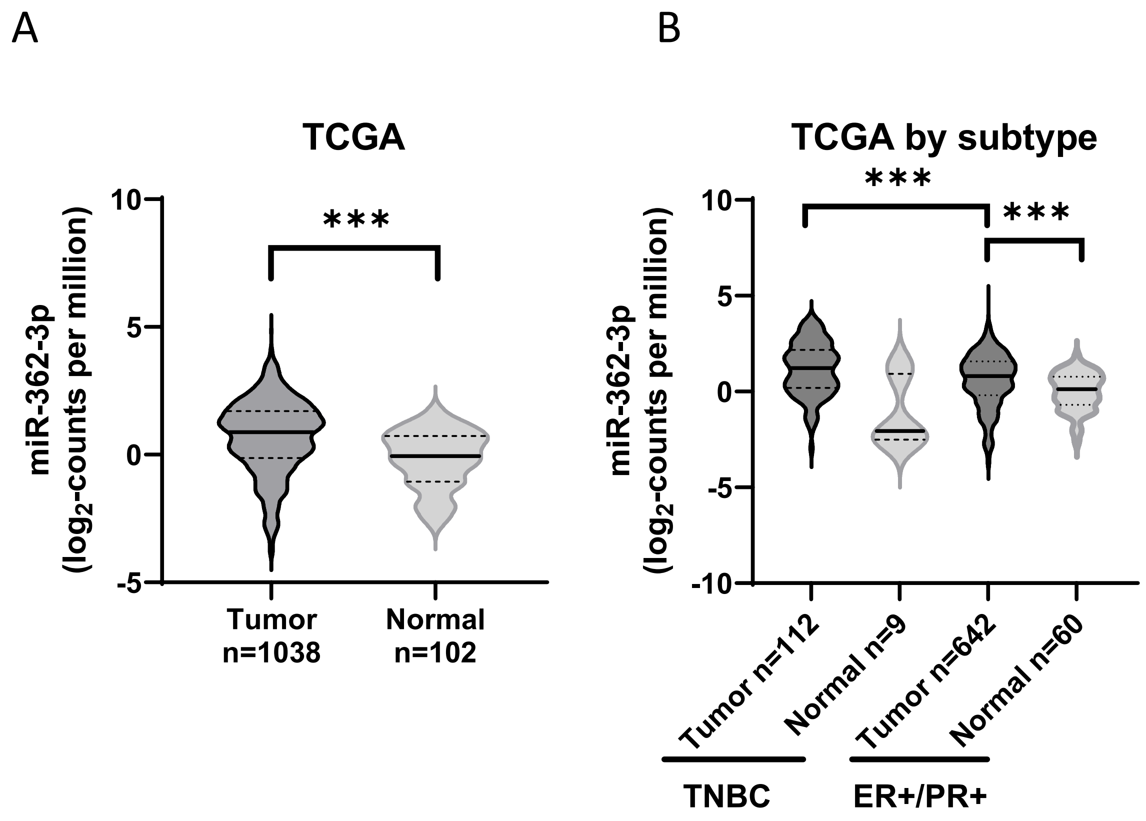


**Fig. S2. MiR-362-3p expression is elevated in breast cancer tissue and enriched in the TNBC subtype**. (**A**) Comparison of miR-362-3p expression levels between breast tumors and adjacent normal breast tissue in the TCGA cohort. (**B**) MiR-362-3p expression stratified by subtype; TNBC tumors had higher miR-362-3p expression than ER+/PR+ tumors. Violin plots indicate the median (solid line) and the 25th and 75th percentiles (dotted lines). *** represents $p<0.001$.

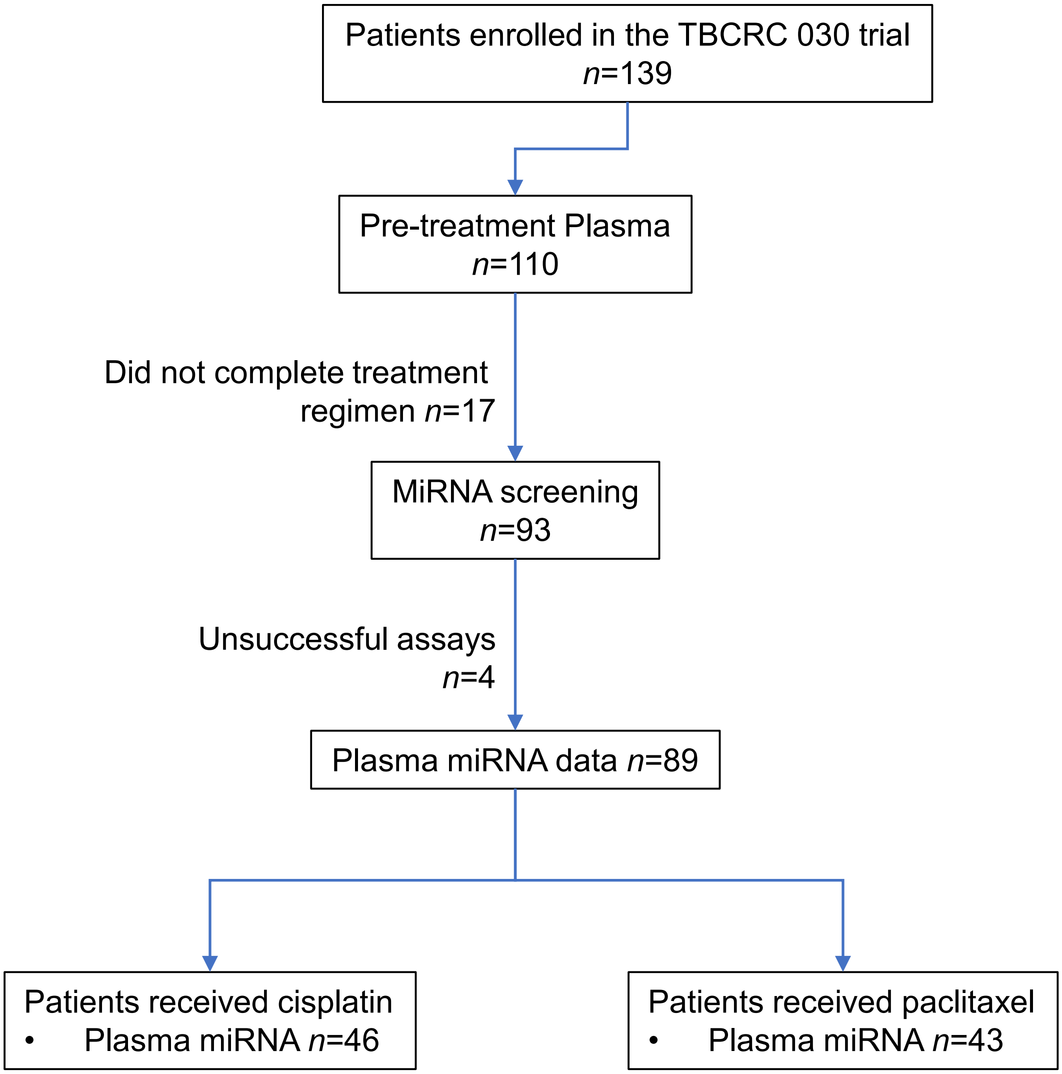


**Fig. S3. Flow diagram of TBCRC 030 patients and miRNA profiling.**

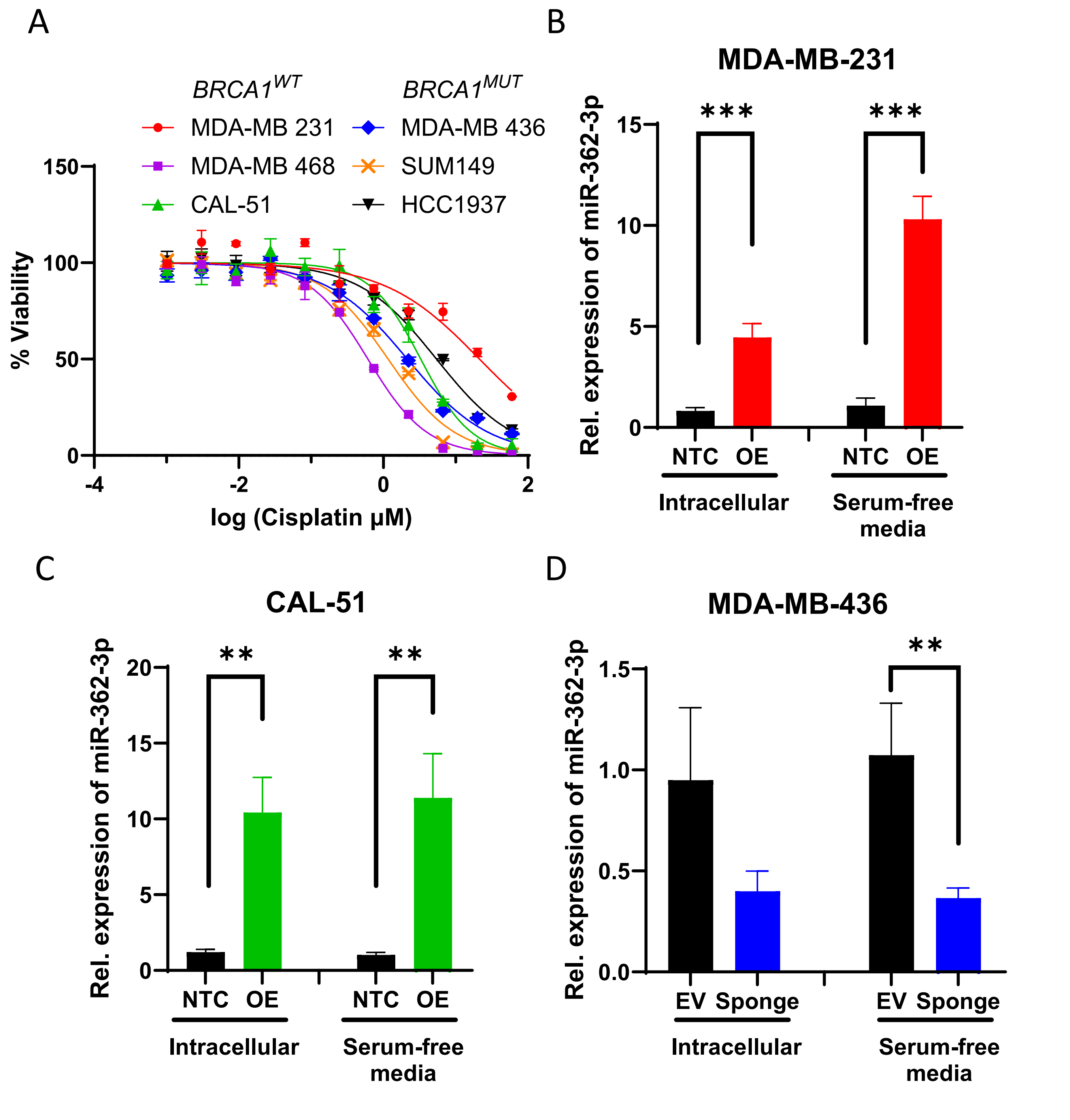


**Fig. S4. Validation of miR-362-3p modulation and baseline cisplatin sensitivity in TNBC cell lines. A.** Baseline cisplatin $IC_{50}$ values in six TNBC cell lines. **B, C.** qRT-PCR shows miR-362-3p levels following stable overexpression (OE) in MDA-MB-231 (**B**) and CAL-51 (**C**) cells compared to non-targeting controls (NTC). **D.** Sponge constructs significantly inhibited miR-362-3p expression in MDA-MB-436 cells relative to empty vector (EV). Data represent mean±SD in bar graphs and mean±SEM in dose response curves. *, ** and *** represent $p$<0.05, $p$<0.01, and $p$<0.001, respectively.

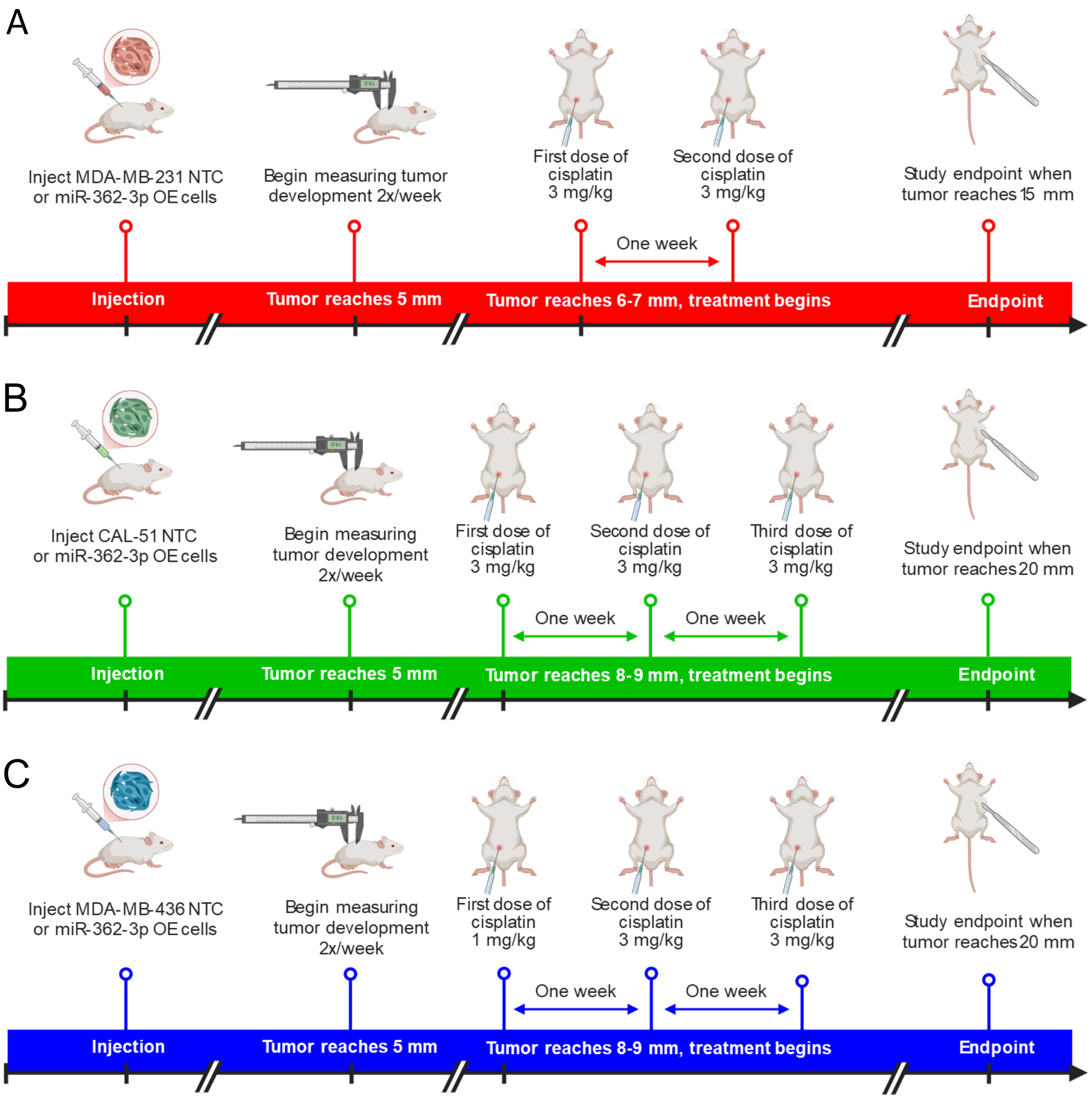


**Fig. S5. Experimental designs of xenograft models.** Cisplatin responses were evaluated in MDA-MB-231 (**A**), CAL-51 (**B**), and MDA-MB-436 (**C**) xenografts. Overexpressing, OE; non-targeting control, NTC; empty vector, EV.

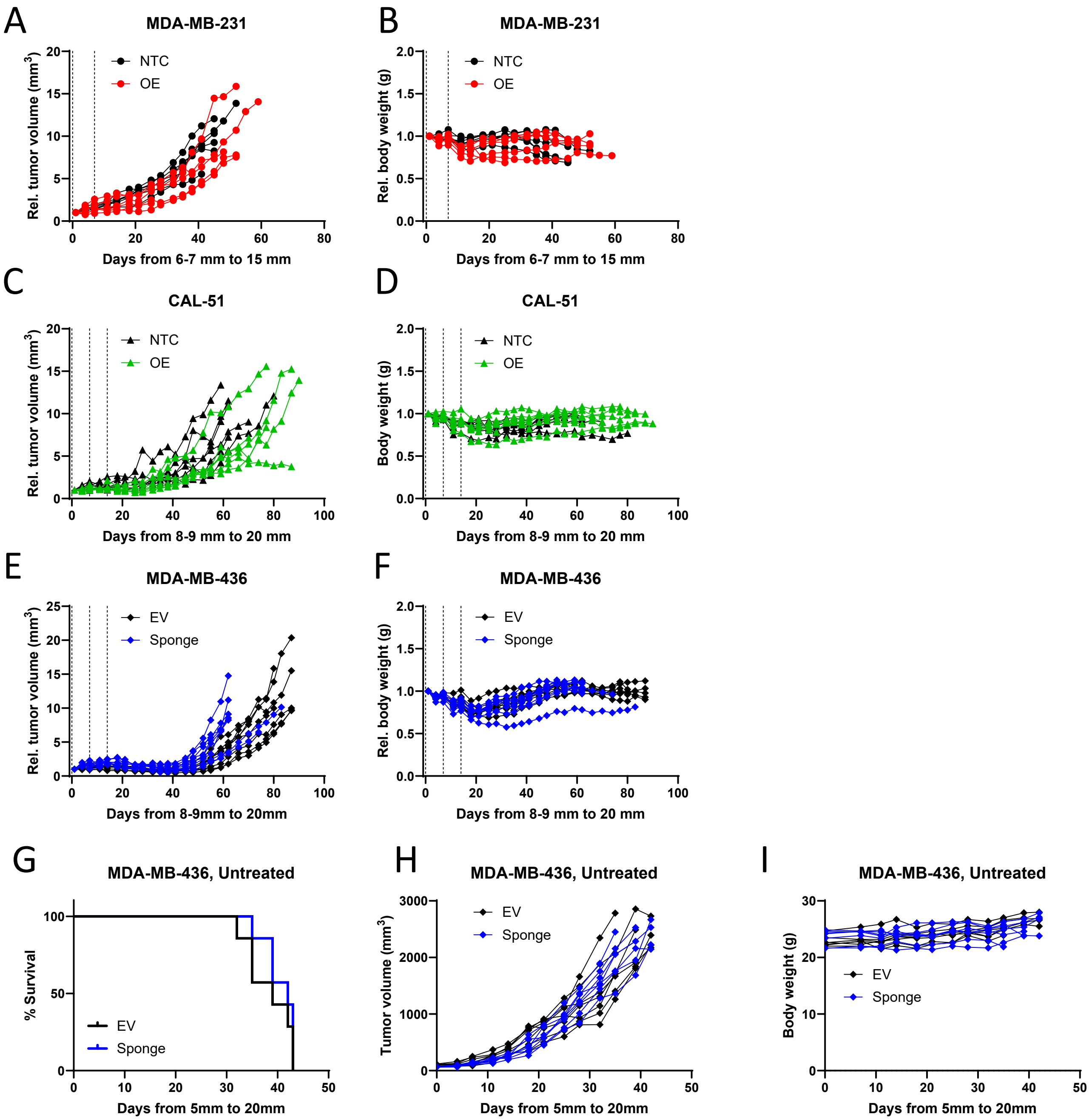


**Fig. S6. Effects of miR-362-3p modulation on tumor growth kinetics and systemic toxicity in TNBC xenografts. A-F.** Longitudinal monitoring of relative tumor volumes and mouse body weights in cisplatin-treated xenografts for MDA-MB-231 (**A, B**), CAL-51 (**C, D**), and MDA-MB-436 (**E, F**) models. Vertical dotted lines indicate the timing of cisplatin administration. **G.** Kaplan-Meier survival analysis of untreated MDA-MB-436 xenografts, demonstrating that miR-362-3p inhibition does not alter baseline tumor-specific survival in the absence of chemotherapy. **H, I.** Longitudinal tumor volumes (**H**) and body weights (**I**) for the untreated MDA-MB-436 cohort. Data represent mean±SEM. Overexpressing, OE; non-targeting control, NTC; empty vector, EV.

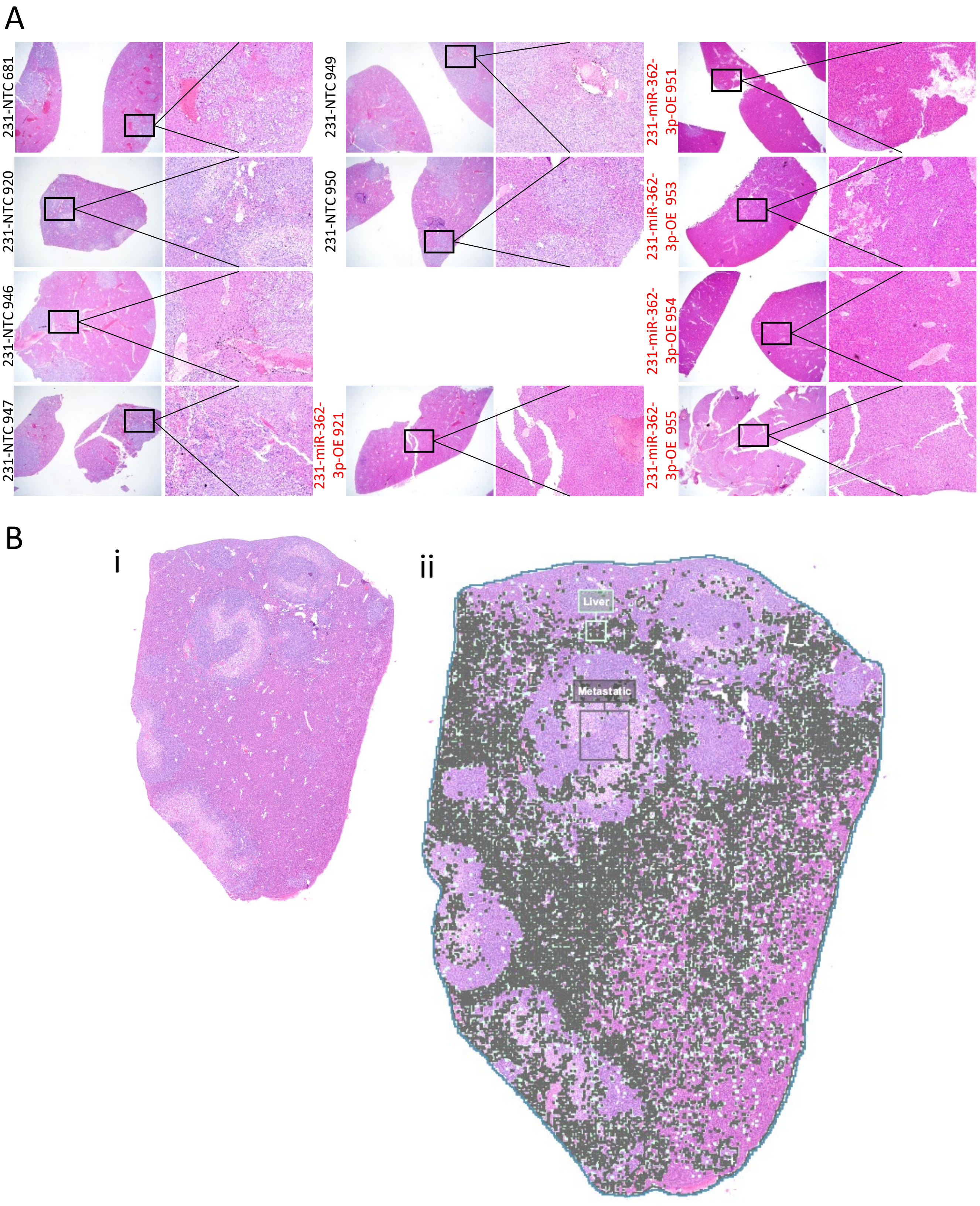


**Fig. S7. Histological assessment of liver metastatic burden. A** H&E-stained liver sections from cisplatin-treated mice bearing MDA-MB-231 xenografts. Left panels provide a wide-field overview at 2×, with 10× high-magnification insets at the right. Overexpressing (OE) miR-362-3p xenografts (red text) exhibit a reduction in metastatic foci compared to non-targeting controls (NTC; black text). **B.** Quantitative analysis of total metastatic burden in liver sections was performed using classification in QuPath. **i.** An example H&E section of the liver. **ii** Pixel classification classified cells as liver (dark green overlay) or metastatic cells.

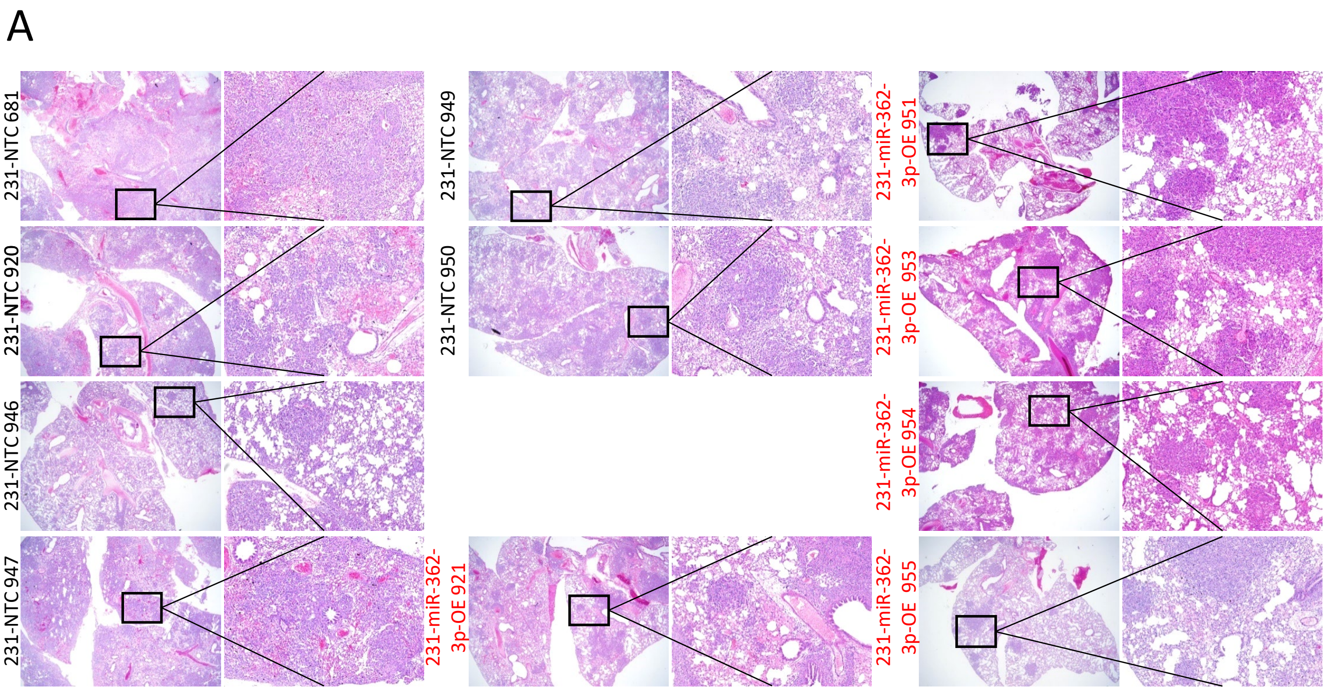


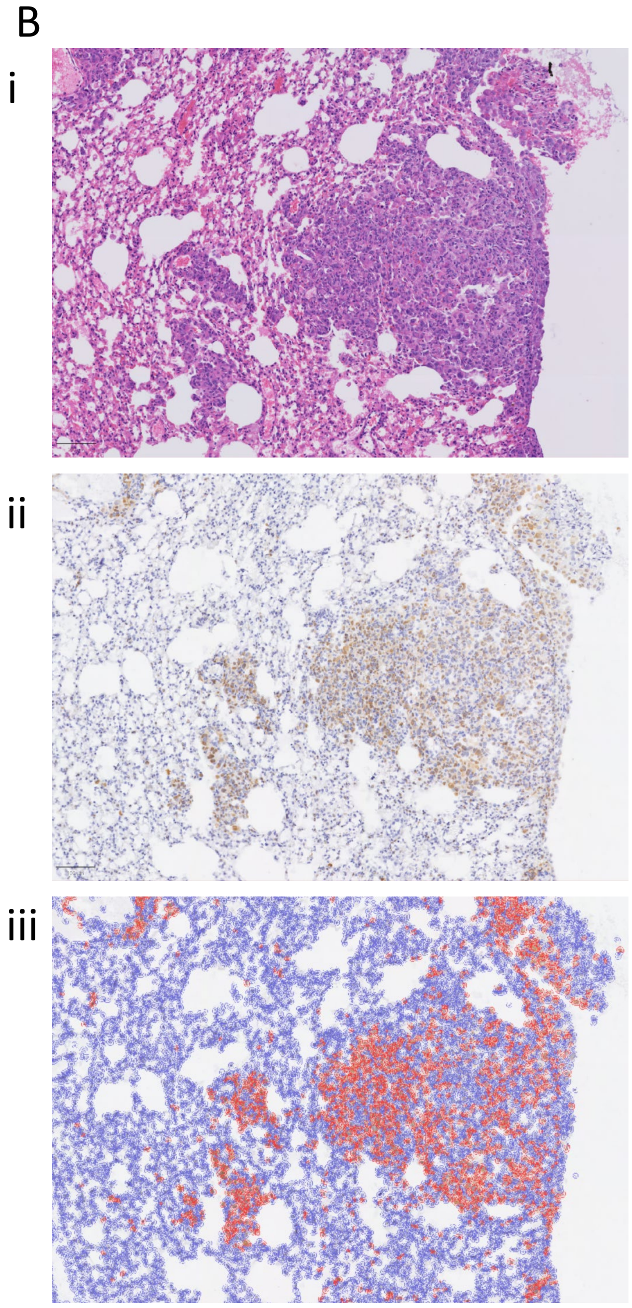


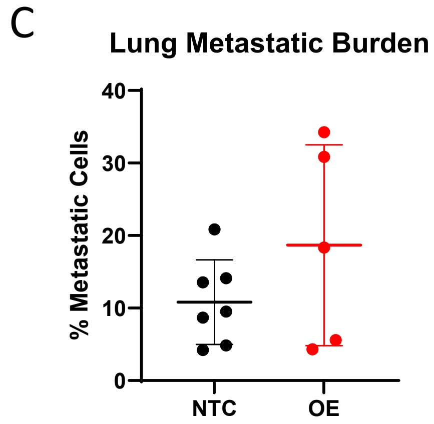


**Fig. S8. Quantitative assessment of metastatic burden in the lungs. A.** H&E-stained lung sections from cisplatin-treated mice bearing MDA-MB-231 xenografts Left panels provide a wide-field overview at 2×, with 10× high-magnification insets at the right. **B.** Example lung H&E image (**i**) with its corresponding IHC section stained with human-specific Ku80 (**ii**; 10×). Image analysis using QuPath to detect cells (blue overlay) and quantify Ku80 positive cells (red overlay). **C.** Quantitative comparison of lung metastatic burden based on Ku80-positive cell counts. No significant difference was observed between miR-362-3p-overexpressing (OE) and non-targeting control (NTC) tissues. Data represent mean±SD.

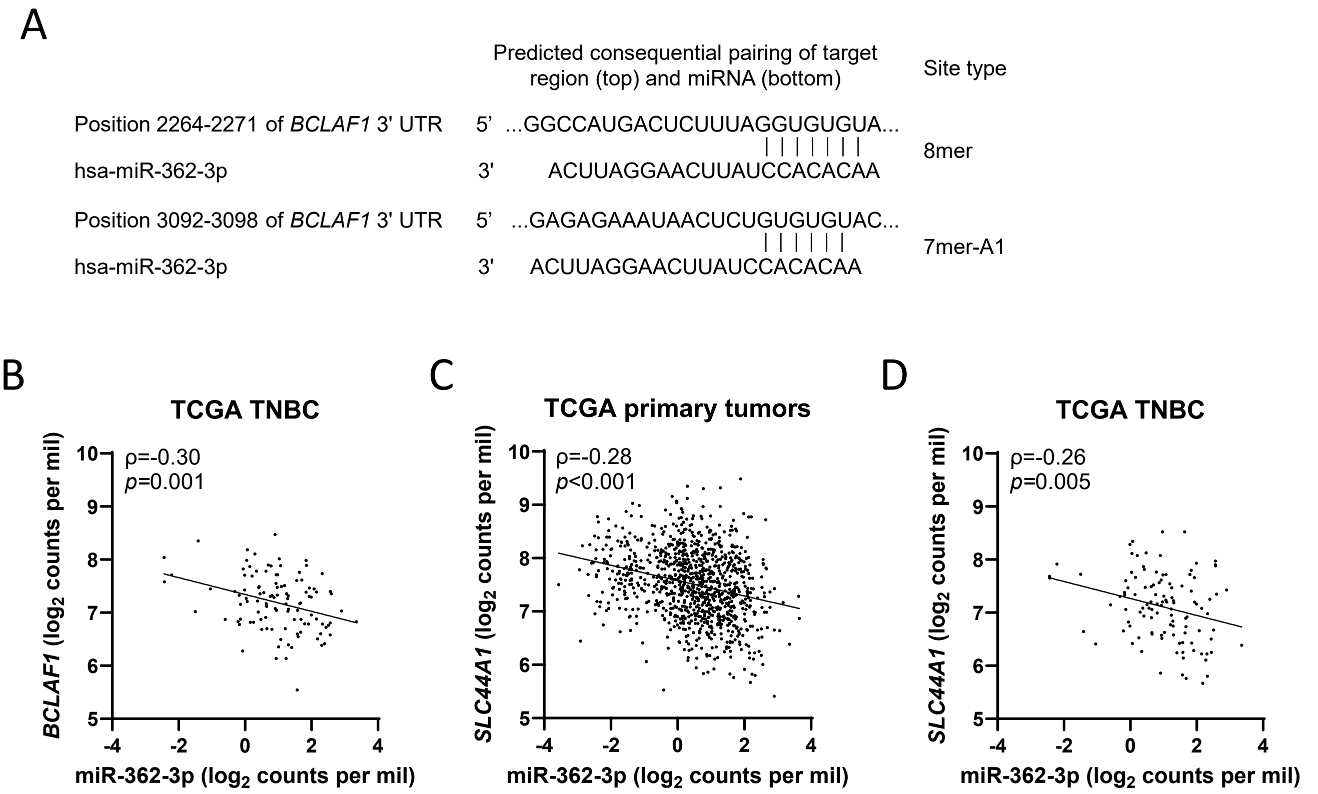


**Fig. S9. MiR-362-3p is predicted to target *BCLAF1* and correlates with *BCLAF1* and *SLC44A1* expression in TCGA. A.** Schematic representation of two conserved miR-362-3p binding sites within the BCLAF1 3' UTR, as predicted by **[???]. B.** Negative correlation between miR-362-3p and *BCLAF1* expression within the TNBC subtype of the TCGA breast cancer cohort. **C-D.** Negative correlations between miR-362-3p and *SLC44A1* expression across all TCGA breast cancer cases (**C**) and the TNBC subset (**D**).

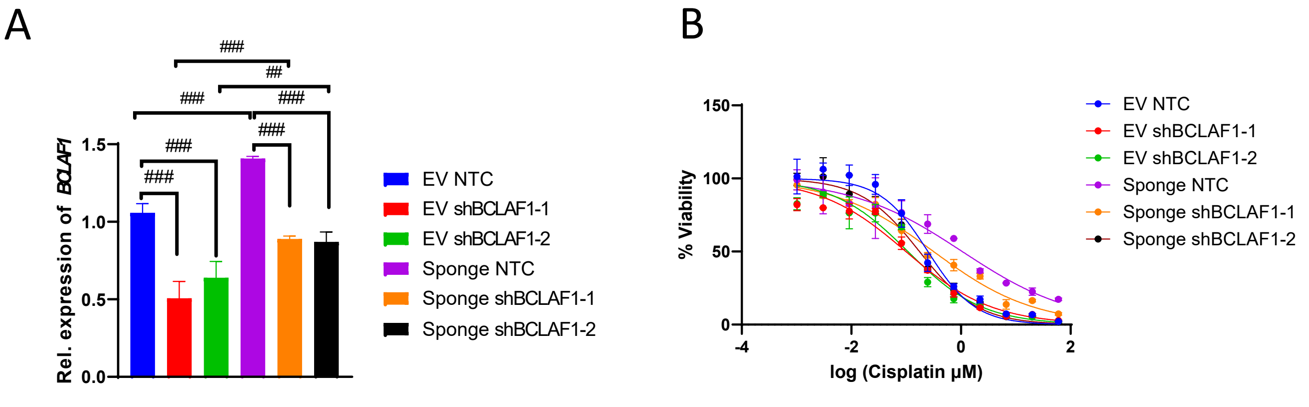


**Fig. S10. Suppression of *BCLAF1* rescues cisplatin sensitivity following miR-362-3p inhibition. A.** qRT-PCR validation of *BCLAF1* silencing in MDA-MB-436 cells utilizing two independent shRNAs (red and green). Inhibition of miR-362-3p significantly induced *BCLAF1* expression (purple), an effect that was effectively neutralized by concomitant shRNA-mediated BCLAF1 knockdown (orange and black). **B.** Cisplatin dose-response curves demonstrating that the chemoresistance phenotype induced by miR-362-3p inhibition (purple) is reversed upon stable suppression of *BCLAF1* (orange and black). Data represent mean±SD in bar graphs and mean±SEM in dose response curves. ## and ### represent adj *p*<0.01 and adj *p*<0.001, respectively. Non-targeting controls, NTC; empty vector, EV.
