## Supplementary material for "The miR-362-3p/*BCLAF1* axis regulates cisplatin sensitivity and metastatic progression in triple-negative breast cancer": Table S2

**Table S2.** Multivariable analysis of miR-362-3p associations with pathologic complete response (pCR; **A**) and residual cancer burden (RCB) score of 0 or 1 **(B)** among 53 cisplatin-treated patients.

|  | **Exp(β)** | **95% CI** | ***p* value** |
| --- | --- | --- | --- |
| A. **pCR** |  |  |  |
| Crude | 3.62 | 1.49-11.0 | 0.01 |
| Minimally adjusted | 4.37 | 1.70-14.5 | 0.006 |
| Fully adjusted | 5.00 | 1.69-22.1 | 0.011 |
| B. **RCB 0/1** |  |  |  |
| Crude | 3.01 | 1.46-7.19 | 0.006 |
| Minimally adjusted | 3.39 | 1.58-8.57 | 0.004 |
| Fully adjusted | 4.63 | 1.72-17.7 | 0.008 |

Minimally adjusted model accounted age at surgery and year of enrollment. Fully adjusted model accounted for age at surgery and year of enrollment, clinical stage, tumor grade, triple-negative breast cancer status, and tumor-infiltrating lymphocytes. Confidence interval, CI.
