## Supplementary material for "The miR-362-3p/*BCLAF1* axis regulates cisplatin sensitivity and metastatic progression in triple-negative breast cancer": Table S3

**Table S4.** Demographic and tumor characteristics of the TBCRC 030 participants, stratified by treatment

|  |  | Overall | Cisplatin | Paclitaxel |
| --- | --- | --- | --- | --- |
| *n* |  | 89 | 46 | 43 |
| Age, years, mean (SD) |  | 51.5 (11.7) | 51.6 (12.9) | 51.5 (10.6) |
| Age, years (%) | <40 | 15 (16.9) | 9 (19.6) | 6 (14.0) |
|  | ≥40 | 73 (82.0) | 36 (78.3) | 37 (86.0) |
|  | Unknown | 1 (1.1) | 1 (2.2) | 0 (0.0) |
| *BRCA^mut^* status, *n* (%) | *BRCA1* | 3 (3.4) | 2 (4.3) | 1 (2.3) |
|  | *BRCA2* | 1 (1.1) | 1 (2.2) | 0 (0.0) |
|  | Wildtype | 76 (85.4) | 40 (87.0) | 36 (83.7) |
|  | Unknown | 9 (10.1) | 3 (6.5) | 6 (14.0) |
| Race, *n* (%) | White | 73 (82.0) | 39 (84.8) | 34 (79.1) |
|  | Black | 7 (7.9) | 3 (6.5) | 4 (9.3) |
|  | Asian | 2 (2.2) | 0 (0.0) | 2 (4.7) |
|  | Mixed | 1 (1.1) | 1 (2.2) | 0 (0.0) |
|  | Other | 3 (3.4) | 3 (6.5) | 0 (0.0) |
|  | Unknown | 3 (3.4) | 0 (0.0) | 3 (7.0) |
| Menopausal status, *n* (%) | Pre | 47 (52.8) | 26 (56.5) | 21 (48.8) |
|  | Post | 42 (47.2) | 20 (43.5) | 22 (51.2) |
| Stage, *n* (%) | I | 10 (11.2) | 6 (13.0) | 4 (9.3) |
|  | II | 66 (74.2) | 33 (71.7) | 33 (76.7) |
|  | III | 13 (14.6) | 7 (15.2) | 6 (14.0) |
| Pre-treatment node status, *n* (%) | Positive | 31 (34.8) | 14 (30.4) | 17 (39.5) |
|  | Negative | 58 (65.2) | 32 (69.6) | 26 (60.5) |
| Histologic type, *n* (%) | IDC | 86 (96.6) | 44 (95.7) | 42 (97.7) |
|  | ILC | 1 (1.1) | 0 (0.0) | 1 (2.3) |
|  | IDC and ILC | 1 (1.1) | 1 (2.2) | 0 (0.0) |
|  | Other | 1 (1.1) | 1 (2.2) | 0 (0.0) |
| Tumor grade, *n* (%) | 2 | 6 (6.7) | 2 (4.3) | 4 (9.3) |
|  | 3 | 83 (93.3) | 44 (95.7) | 39 (90.7) |
| pCR, *n* (%) | Yes | 12 (13.5) | 6 (13.0) | 6 (14.0) |
|  | No | 77 (86.5) | 40 (87.0) | 37 (86.0) |
| RCB, *n* (%) | 0/1 | 22 (24.7) | 11 (23.9) | 11 (25.6) |
|  | 2/3 | 67 (75.3) | 35 (76.1) | 32 (74.4) |

SD, standard deviation; IDC or ILC, invasive ductal or lobular carcinoma; pCR, pathologic complete response; RCB, residual cancer burden.
